# Learning diverse and generic representations of the brain with large-scale multi-task pretraining

**DOI:** 10.64898/2025.12.19.25342659

**Authors:** Esten H. Leonardsen, Andreas Dahl, Madelene C. Holm, Ann-Marie de Lange, Edvard O. S. Grødem, Andre F. Marquand, Øystein Sørensen, Emanuel Schwarz, Ole A. Andreassen, Thomas Wolfers, Yunpeng Wang, Lars T. Westlye

## Abstract

Large pretrained models developed and shared by actors with privileged access to data and compute have played a central role in the democratisation of deep learning in a range of domains. Here, we contribute to this endeavour in the field of neuroimaging, by compiling a large dataset of structural magnetic resonance imaging scans (n=114,257) and using them to pretrain a multi-task convolutional neural network to predict age, sex, handedness, BMI, fluid intelligence and neuroticism. Subsequent analyses show that our pretraining approach results in a rich and diverse feature space, implying the model is sensitive to a broad spectrum of nuanced neuroanatomical variation. We also demonstrate the usefulness of the pretrained model by employing it to predict brain age and perform clinical classifications via transfer learning, showing that it consistently outperforms models trained from scratch.

## Introduction

Recent years have seen the broad adoption of artificial intelligence and deep neural networks (DNNs) for predictive tasks in a variety of domains, often shattering existing performance benchmarks (LeCun et al., 2015). This has in part been made possible by the democratisation of knowledge and technology, with key theoretical insights shared through scientific dissemination and implemented in efficient, open-source frameworks, enabling their use by a vast community of technologists (Zhou et al., 2024). Still, training state-of-the-art DNNs requires quantities of data and computational resources that are inaccessible to many (Ahmed & Wahed, 2020). To alleviate this problem, the deep learning community has adopted and advanced a culture of sharing pretrained models, typically large and complex DNNs trained on immense amounts of data to solve generic predictive problems. These models have subsequently been employed to solve new tasks through the process of transfer learning (Bengio, 2012), which can lead to good performance even when training data are limited (Weiss et al., 2016). Here, a pretrained model comprises a starting point for further training towards a new objective, under the assumption that knowledge attained by the model to solve the original problem is somehow useful towards the downstream task. Notable examples are convolutional neural networks (CNNs) pretrained on the ImageNet database (Deng et al., 2009) that have been successfully fine-tuned towards a diverse array of visual predictive tasks (Morid et al., 2021), playing a central role in the image processing revolution of the last decade (Huh et al., 2016).

Also in clinical neuroscience, deep learning models, often CNNs, are becoming attractive alternatives for processing neuroimaging data (Ebrahimighahnavieh et al., 2020; Quaak et al., 2021). CNNs have been trained on structural magnetic resonance imaging (MRI) data to differentiate healthy controls (HCs) from patients with a variety of clinical conditions (Quaak et al., 2021), predict clinical outcomes (Janssen et al., 2018), and derive generalised markers of brain health (Cole et al., 2017). Yet, the adoption of these methods is impeded as most neuroimaging datasets are relatively small, consisting of hundreds or thousands of participants (Madan, 2022), orders of magnitude smaller than ImageNet and its counterparts. This issue is particularly pronounced in clinical datasets, containing variables constituting clinically relevant prediction targets, that are often substantially smaller than population datasets comprised of relatively healthy participants. Motivated by this predicament, transfer learning is becoming a popular alternative also in clinical neuroimaging studies (Valverde et al., 2021).

Transfer learning presupposes the existence of suitable pretrained models, whose construction relies on several design choices known to impact their down-stream utility. Among these, the arguably most important is the selection of a pretraining objective. In neuroimaging studies employing transfer learning, pretraining has most commonly been supervised (Agarwal et al., 2021), by employing the same predictive task that will be used downstream (Holderrieth et al., 2023), or learning to predict a generic target variable that occurs across multiple datasets, such as age (Bashyam et al., 2020; Leonardsen et al., 2022) or sex (B. Lu et al., 2022). However, recent work indicates that pretraining using surrogate markers such as age offers little advantage, even when the pretraining target is presumably related to the downstream task (Tan et al., 2025). In constrast, the broader computer vision community has strived for generality during pretraining, exemplified in the ImageNet case where pretraining entails learning to recognise images from thousands of diverse classes. Still, even this very broad classification scheme has shown limited transferability to narrow and specialised downstream tasks (Kornblith et al., 2019), and the most prevalent pretraining approaches are self-supervised. Here, pretraining does not rely on preexisting labels, but rather detecting automatically inferred targets (Caron et al., 2019), reconstructing input data (K. He et al., 2021), or understanding semantic similarities (Chen et al., 2020), all with the goal of robustly learning to detect important features from the input space (Bengio, 2012).

Despite their popularity, self-supervised pretraining methods are known to exaggerate the importance of salient patterns in the pretraining data while overlooking nuances (Chen et al., 2020; Li et al., 2021), plausibly causing detriment when transferring to tasks relying on subtle morphological differences in structural neuroimaging data. Between self-supervision and single-task supervised approaches, multi-task learning (Caruana, 1997) represents a promising alternative to guide models during pretraining towards patterns that can subsequently be useful across a broader spectrum of downstream tasks (Baxter, 2000). This is achieved by optimising several, often supervised, objectives simultaneously, leading the model to learn features that are inherently general, while demonstrably being predictive for a given set of targets. In neuroimaging data, the multi-task approach represents a promising venue to discover variability among healthy individuals that generalises to patient groups, potentially bridging the gap between large population datasets and smaller clinical samples. Nonetheless, the utility of multi-task learning for MRI-based transfer learning remains largely unexplored.

In this work, we pretrain a state-of-the-art CNN architecture on a large, composite dataset of structural MRI scans. By combining architectural alterations and a multi-task pretraining regime, we argue that this model constitutes a generalised backbone for neuroimaging data, useful for transfer learning towards specialised downstream tasks. We demonstrate that the feature space encoded by the multi-task model exhibits more diversity than a commonly used single-task pretraining alternative, implying that it has learned to detect and leverage a broader set of morphological patterns in the scans. Finally, we showcase the utility of the model by applying it to two downstream age prediction tasks and two clinical classification tasks. Both the code and the weights of the pretrained model and accessible interfaces for both inference and transfer learning are available on GitHub, HuggingFace and DockerHub.

## Methods

### Data

The data used in this work was obtained from previously published studies approved by their respective institutional review boards or relevant research ethics committees (Supplementary Table 1). The age and sex distribution of the full dataset (114,257 images from 80,931 participants, mean age=50 years±26 years standard deviation, 49% females, originating from 45 unique cohorts) can be seen in Figure 1. A per-sample overview of the number of participants and images, age and sex distributions, and their role in the modelling process can be found in Supplementary Table 2.

**Figure 1.**
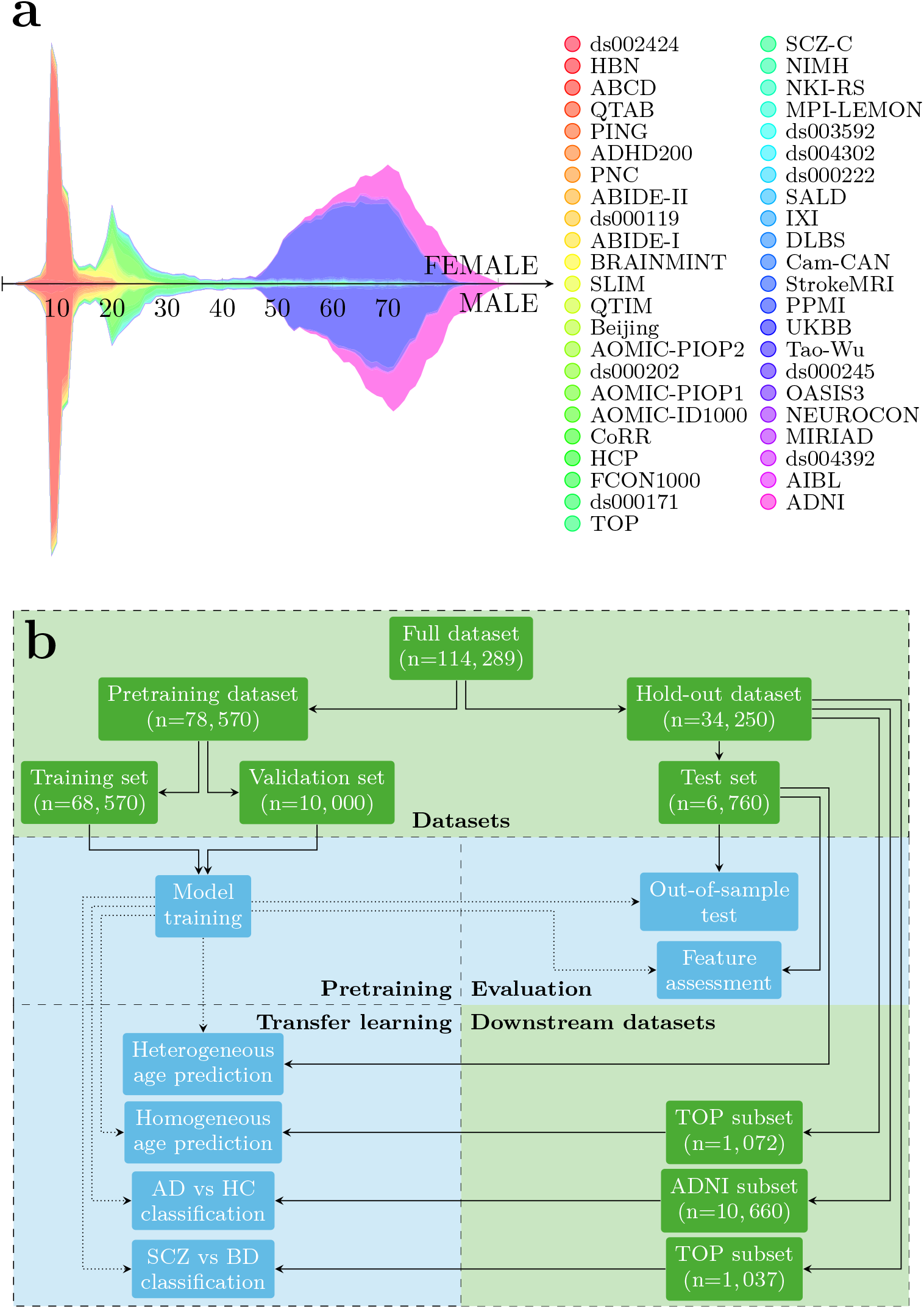
Overview of the data and modelling pipeline in the study. (a) Age and sex distribution in the full dataset. (b) Schematic of data partitioning, modelling and evaluation tasks. Solid lines indicate data-flow, dotted lines indicate propagation of a pretrained model. TOP=Thematically Organized Psychosis dataset, ADNI=Alzheimer’s Disease Neuroimaging Initiative dataset

For pretraining models we used 78,570 brain scans from 69,923 unique, healthy, participants, originating from 26 cohorts (mean age=46±25 years, 51% females). While stratifying on source, age, and sex, this dataset was split into a training (68,570 images, 59,923 participants, mean age=46±26 years, 51% females, Supplementary Figure 1a) and validation (10,000 participants with a single image each, mean age=49±24 years, 52% females, Supplementary Figure 1b) fold, ensuring that all images from a given participant resided solely in one of the two folds.

For testing the predictive performance of the models and evaluate their utility in downstream tasks, we compiled a held-out test set consisting of 34,250 images from 13,442 participants (mean age=60±24 years, 45% females), originating from 20 cohorts not seen during pretraining. The only exception to this rule was data originating from Alzheimer’s Disease Neuroimaging Initiative (ADNI), where we reserved the oldest healthy participants (age ≥ 90 years) for pretraining to ensure the models were trained with an age range covering that of the test set. The remaining ADNI participants (age *<* 90 years) were reserved for testing and downstream tasks.

The held-out data were used for multiple purposes. First, we used it to calculate unbiased results for the predictive tasks employed during pretraining, described in detail below. To this end, for all participants that at some point in time were considered healthy, we extracted a single image from the latest timepoint without a diagnosis. In total, this yielded a test set of 6,760 images from the same number of participants (mean age=38±26 years, 48% females, Supplementary Figure 1c).

In the four downstream tasks (described in detail below), we sampled datasets of varying sizes from an overarching pool constructed specifically for each task. For the first age prediction task this pool consisted of the same 6,760 images that were used as a test set above, representing a heterogeneous population originating from a multitude of cohorts and spanning a wide age range. For the second age prediction task, the pool contained 1,072 images from the same number of healthy participants in the Thematically Organized Psychosis (TOP) cohort (mean age=34±10 years, 46% females), representing a more homogeneous population from a limited number of scanners with a narrower age range. For a task concerned with differentiating patients with Alzheimer’s disease (AD) and HCs, the pool comprised a subset of participants from ADNI (10,660 images from 1,678 participants, 718 with AD, 960 HCs, mean age 75±7 years, 50% females). For a task concerned with differentially diagnosing patients with schizophrenia (SCZ) and bipolar disorder (BD), we used a subset of participants from TOP (1,037 images from 861 participants, 473 with SCZ, 388 with BD, mean age=32±10 years, 47% females).

As input to all models, we used T1-weighted structural MRIs. Prior to modelling, these were minimally preprocessed to ensure they were spatially aligned and contained minimal amounts of non-brain tissue. This was achieved using FastSurfer (Henschel et al., 2020) version 2.0.1, that rotates the image to RAS orientation and produces a binary brainmask. The finalized input was procured by multiplying the rotated image with the mask to retain only brain tissue, and cropping a 224×192×224 cube centered around the brain to remove empty space.

To accompany the images used as inputs during pretraining we compiled two sets of predictive targets to act as outputs, corresponding to two distinct pretraining strategies. The first contained only age, such that the pretraining process resulted in a standard brain age model. For the second, we constructed a vector with six entries per image: age, sex, handedness, body mass index (BMI), fluid intelligence (Supplementary Table 3), and neuroticism (Supplementary Table 4), such that the pretraining yielded a multi-task model. These specific variables were selected pragmatically, representing a diverse set of demographic, physical, and psychological characteristics, that were programatically retrieveable from the larger cohorts and had limited correlation (Supplementary Figure 5). To train the model, age was retained as-is, with a single continuous number per timepoint encoding the chronological age of the participant at the time of the scan (min age=3, max age=97.4, no missing entries in the training set). Sex and handedness were coded as binary variables (female=0, male=1, 1 out of 68,570 participants had a missing entry; right-handed=0, left-handed=1, 48,545/68,570 missing entries). BMI was retained as an unmodified continuous variable (mean=24.53±5.18, 17,516/68,570 missing). Both fluid intelligence (24,615/68,570 missing) and neuroticism (37,332/68,570 missing) denoted different measurements in the various data sources, and, to unify them into a singular target with approximately the same meaning, each variable was z-score standardized within each independent data source before they were merged. Finally, these six variables were stacked into a target vector **y** = [*y*_0_, *y*_1_, *y*_2_, *y*_3_, *y*_4_, *y*_5_] for each image, where missing values were retained as NAs to be handled later in the process.

### Model architectures

To match the two pretraining strategies we trained two types of models, both relying on the Simple Fully Convolutional Network (SFCN) architecture (Gong et al., 2021; Peng et al., 2021) as a backbone. This is a relatively simple CNN, starting with five repeated blocks consisting of a 3-dimensional convolution (3×3×3 filters), batch normalisation, a rectified linear unit (ReLU) activation, and a 3-dimensional max pooling layer (2×2×2 patches). After these blocks comes another 3-dimensional convolution (1×1×1 filters) for downsampling, batch normalisation, and a final ReLU activation. At this point, the architecture of the two models diverged. The first model predicting only age was a regression variant of SFCN (SFCN-reg), a brain age model that has previously been shown to surpass other variants in terms of out-of-sample generalisation (Leonardsen et al., 2022). In this model, the last ReLU was succeeded by a global average pooling layer followed by a dropout layer. Finally came the prediction layer, with a single output neuron with a linear activation, allowing for a singular, continuous, brain age prediction per input image. In the multi-task model (SFCN-multi), after the last ReLU in the backbone, we instead employed a global max pooling layer, under the assumption that this would produce a more diverse feature space. Here, we also removed the dropout layer, due to its propensity to increase feature redundancy, and the innate regularizing effect of multi-task training (Cao et al., 2019). Finally, SFCN-multi had a prediction head with six output neurons to match the entries of the multi-task target vectors. The first, fourth, fifth, and sixth artificial neurons, corresponding to age, BMI, fluid intelligence, and neuroticism respectively, were neurons with a linear activation, facilitating the prediction of a continuous target. The second and third, corresponding to sex and handedness, had a sigmoid activation to enable binary predictions. During training, the loss for SFCN-multi was aggregated across all of these outputs.

### Training

Both models were implemented and trained using TensorFlow version 2.6 (Abadi et al., 2015) distributed across 4 Nvidia A100 GPUs. Each of these had 40GB VRAM, incurring ∼2-3 days of training per model. For both SFCN-reg and SFCN-multi, we fit a single model instance with a fixed set of hyperparameters based on results from an earlier study (Leonardsen et al., 2022), i.e. a weight decay of 10^*-*3^, previously observed to yield the best results across multiple architectural variants, and, for SFCN-reg, a dropout rate of 0.2, which previously produced the best models of this type. Both models were fit with an Adam optimizer and an initial learning rate of 10^*-*3^ that was reduced in a stepwise manner by a factor of 10 at epochs 30 and 50 (out of a total of 60). After training, weights were restored from the epoch yielding the lowest loss in the validation set.

SFCN-reg, predicting only age, was trained to minimize the mean squared error (MSE) of the age predictions. SFCN-multi, with its six output neurons, employed different loss functions *ℓ*_0_,...,*ℓ*_5_ for each neuron: an MSE-loss for the first, fourth, fifth and sixth output neuron corresponding to regression tasks, and binary cross-entropy (BCE) for the second and third, corresponding to binary classifications. Intuitively, the composite loss *L* used for fitting the model should be computed by summing over these task-specific losses, 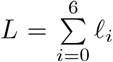. However, if left unchecked, the exponential nature of MSE and the dampening effect of BCE, combined with varying magnitudes of the target variables, would produce losses of different scales, effectively prioritizing some tasks over others (Cao et al., 2024). To alleviate this, we employed a pragmatic, two-step heuristic approach to find a weighting scheme that strove for losses approximately on the same scale, inspired by adaptive methods (Kendall et al., 2018). In the first step, we aimed to normalise the loss scales without regard for task difficulty or input data. To achieve this, we calculated the relative scale between losses procured by a dummy multi-task model

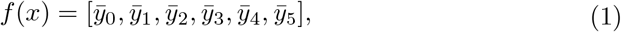

where 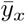 refers to the mean of target variable *y* in the training set.

More precisely, we calculated the mean loss 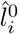 for each output-variable *y*_*i*_ in the training set based on the predictions from this model, and used their inverses as an intermediate set of weights

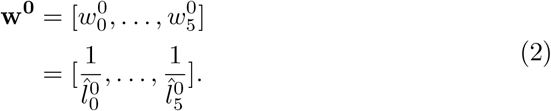

For the second step, we aimed to incorporate a term that adjusted for task difficulty conditioned on the input data, after eliminating differences in magnitude using **w**^**0**^. This was achieved by fitting a CNN optimizing an intermediate, weighted loss

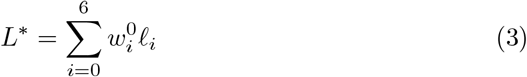

for five epochs. Then, we noted the mean 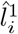 of each of the individual weighted losses 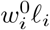 across all batches in the fifth epoch, indicating the relative difficulty the model had with each task at that point in the training process. This gave rise to a new set of weights

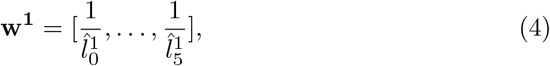

conditioned upon **w**^**0**^. The final set of weights was computed as the element-wise product of these two weight vectors,

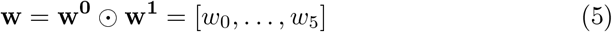

and was used to calculate the final, weighted composite loss 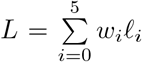 employed during training. In addition to the task weights, we employed class-specific weights for the binary classification tasks to alleviate the effects of class imbalance, by assigning a weight proportional to the inverse of its frequency to each class (King & Zeng, 2001). For each image missing the ground truth value for one or more of the six target variables, the losses from the corresponding tasks were masked out.

### Evaluation

We assessed the performance of the models using both the validation set and the held-out test data. The age predictions from both SFCN-reg and SFCN-multi were evaluated by calculating their mean absolute error (MAE), and the same measure was used to evaluate the BMI predictions of SFCN-multi. For the sex and handedness-predictions, both binary tasks, we evaluated performance by calculating the area under the receiver operating characteristic curve (AUC). For fluid intelligence and neuroticism, we calculated Pearson’s R to quantify performance. For the predictions relying on AUC and R we relied on theoretically derived baseline measures to represent a comparative basis (e.g. AUC=0.5, R=0.0) for interpretation of predictive performance. For age, we contextualized performance by pitting the MAEs of the SFCN-reg and SFCN-multi against each other. For BMI, we constructed a baseline by computing the MAE from a very simplistic model generating its predictions stochastically by mirroring a normal distribution with mean and standard deviation derived from the ground truth data.

To evaluate feature diversity, we extracted feature vectors from the second-to-last (bottleneck) layer of both SFCN-reg and SFCN-multi, based on the 6,760 participants in the held-out test set. With these vectors we performed two correlational analyses, both relying on the Pearson correlation coefficient. First, we calculated and visualised the intercorrelation between the features for qualitative interpretation, and aggregated them into a single quantity by taking their mean. Second, we computed the correlation between each of these features and 96 regional volumes derived using FastSurfer. Here, we also calculated the mean of the cosine distances

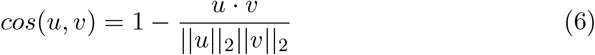

between each pair of feature-wise correlation vectors (i.e. the vector of correlations between a single feature and all regions) *u* and *v* to quantify their difference with regards to global volumetric patterns.

### Transfer learning

To assess the potential benefit of using the pretrained models in novel contexts, we fit tens of thousands of new models to solve four downstream predictive tasks. First, to investigate their usefulness for predicting targets already present in the multi-task scheme in smaller datasets, we fit new models to predict age in two distinct downstream datasets. As a third downstream task, we trained binary classifiers to differentiate between patients with AD and HCs. Here, the boot-strapped samples were drawn balanced to AD patients and HCs by including the same number of subjects from both groups matched stochastically on age and sex. For the fourth downstream task, we trained classifiers to differentially diagnose patients with SCZ and BD by drawing bootstrap samples while matching the two patient cohorts with the same procedure as for the AD vs HC classifiers.

In addition to varying with respect to predictive targets, the models trained for downstream tasks employed different transfer learning strategies. For the age models, where the pretrained model already contained a relevant prediction head, the first strategy generated predictions by keeping the model as-is without further training (referred to as “transferred” models below). In addition to these, we retrained models for the downstream tasks, either by keeping the pretrained model as a fixed feature extractor by freezing the weights in all but the last layer (a strategy yielding models denoted as “frozen”), or using the pretrained weights as initialisation before retraining all layers (referred to as “finetuned” models). For the downstream age models, using the frozen and finetuned strategies, we initialized all layers, including the prediction head, using weights from the pretrained model. For the two predictive tasks that did not exist in the multi-task pretraining scheme (e.g. AD vs HC and SCZ vs BD classification) we recreated a prediction head with random initialisation and a sigmoid activation. Taken together, these three strategies for transfer learning relying on a pretrained model are collectively referred to as “pretrained” models below. Finally, to have a baseline to evaluate their performance against, we trained specialised models for each downstream task from scratch, with the exact same architecture (referred to as “baseline” models) using the default Glorot Uniform initializer in Tensorflow for initialisation.

Besides having different predictive targets and pretraining strategies, the downstream models varied with respect to the number of samples used to train and test them, to investigate whether the optimal transfer learning strategy varied as a function of dataset size. For the homogeneous age task, based on the pool with the most held-out data available, we emulated dataset sizes *n* ∈ {100, 250, 500, 1000, 2500}. In the remaining tasks we emulated *n* ∈ {100, 200, 300, 400, 500}. Given an *n*, we bootstrapped a training set consisting of *n* samples with replacement. To ensure some natural variation with regards to the predictive target in the two age prediction tasks, we enforced a minimum age span of 20 years in each bootstrapped training set. Next, after removing all participants used for training, and constricting the remaining pool to cohorts used for training with participants falling within the age-range of the training set, a validation set of 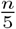 samples was drawn without replacement. And finally, after removing the participants used for validation from the pool, a test set of 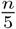 samples was drawn without replacement.

To give an estimate of uncertainty, we bootstrapped 50 triplets of training, validation, and test sets for each specific combination of a predictive task, a transfer learning strategy, and a given *n*. Based on these bootstrapped samples, we trained new models. All of these were trained for 100 epochs, with an initial learning rate of 10^*-*3^ that was annealed by a factor of 10 after not seeing improvement in the validation loss (MSE for regression models, BCE for binary classifiers) for 5 epochs. The training process was early-stopped after 10 epochs without improving the validation loss, and the weights from the epoch yielding the lowest loss in the corresponding validation set was restored. To simulate a realistic setting for transfer learning, we performed a simple grid-based hyper-parameter search for each bootstrapped dataset, over 9 regularisation settings (dropout rate ∈ {0.1, 0.3, 0.5}, weight decay ∈ {1*e*^*-*2^, 1*e*^*-*3^, 1*e*^*-*4^}), and selected the model yielding the lowest validation loss. Finally, this model was applied to the corresponding test set to assess out-of-sample predictive performance. Here, we calculated the Relative Absolute Error,

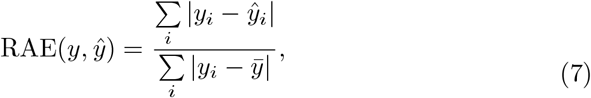

where 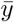 refers to the mean across all labels *y*_*i*_, for the age prediction tasks, to account for age differences in the bootstrapped samples, and AUC for the binary classification tasks. In total, the four down-stream tasks, five unique dataset sizes, 50 bootstrapped samples, three or four transfer learning strategies (depending on whether a direct transfer was possible) and nine hyperparameter settings yielded a total of 29,250 models trained for downstream tasks using these strategies.

To contextualise the performance of the pretrained multi-task model further, we fit additional models for the two downstream classification tasks. First, we fit linear models using the same bootstrapped samples to assess the overall aptitude of deep learning. These were logistic regression classifiers implemented in scikit-learn (Pedregosa et al., 2011) version 1.5 with an *l*_1_-regulariser. As input to these models we used volumes of 96 brain regions, both cortical and sub-cortical, derived from the FastSurfer segmentation maps. To match the multi-task model selection scheme, we fit 9 linear classifiers for each bootstrapped training sample using different regularisation parameters (λ ∈ {1*e*^*-*4^, 1*e*^*-*3^,..., 1*e*^4^}), and selected the model yielding the lowest loss in the corresponding validation set, before generating out-of-sample predictions in the corresponding test set. Next, we wanted to investigate the transfer learning dynamics across single-task and multi-task learning specifically. Motivated by the well-established association between brain age and AD (Franke & Gaser, 2012), we finetuned SFCN-reg towards the AD vs HC classification task, employing the exact same setup as for SFCN-multi.

To quantify the relative efficacy of the different modelling approaches we performed pairwise comparisons between all strategies. For each unique combination of a downstream task, sample size, and strategy *S*, we compiled a list of the 50 out-of-sample performances (RAEs for the age prediction tasks and AUCs for the binary classification tasks),

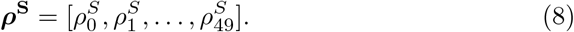

For each pair of strategies *S*_0_ and *S*_1_, where *S*_0_ had superior performance (i.e. 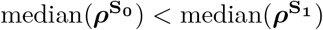 for RAEs, and the opposite for AUCs), we formed the paired list

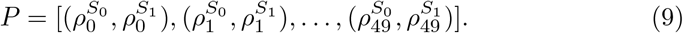

We then computed the median difference between entries in the pairs in this list,

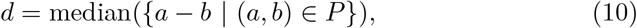

capturing the median performance gain of strategy *S*_0_ over *S*_1_. Next, we compiled 10,000 permuted variants of *P* by shuffling its contents in a pair-wise fashion. For each of the 10,000 iterations, we randomly sampled 50 booleans

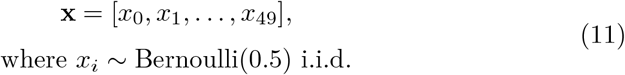

and generated a permutation *P*′ by swapping the internal order in each pair in *P* according to this boolean list,

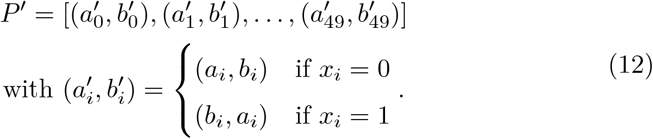

Next, a median was calculated across this permuted list of pairs,

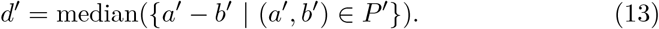

The set of 10,000 permuted median differences *d*′ comprised the null distribution *D*′. Finally, we calculated a one-sided p-value indicating whether the true median difference *d* reflected a significant improvement over *D*′ by computing the proportion of samples in *D*′ with better performance (lower for RAEs, higher for AUCs),

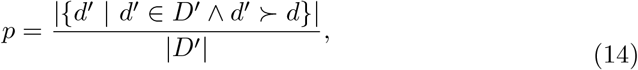

where ≻ indicates better performance and |·| represents the size of ·.

### Pretraining target ablation study

Finally, we investigated the importance of specific variables in the pretraining scheme. Due to its established association with various other phenotypes (Kaufmann et al., 2019; Leonardsen et al., 2022), age was hypothesized to be the most impactful variable among the set of pretraining targets. Thus, we performed an ablation study where we pretrained two models while removing the effect of age in minimally invasive ways. First, by simply removing age from the multi-task scheme, retaining the five other targets as they were. Second, by pretraining across all six pretraining targets (i.e. also retaining age), but scrambling the age labels, effectively making this an impossible target to learn for the model. Next, we compared transfer learning from these two ablated models with both the baseline models and transfer learning from SFCN-reg and SFCN-multi for the two downstream classification tasks. For both tasks we performed the comparison at *n* = 300, the midpoint in terms of sample size. We first investigated whether finetuning the ablated models outperformed the baseline model using the same one-sided permutation test as for the model comparison above. Then, we assessed whether the finetuned ablated models yielded results significantly different from finetuning the brain age model using a two-tailed version of the same test, with

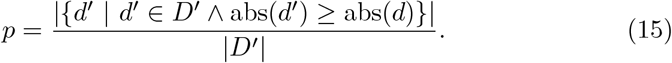

And lastly, we calculated whether finetuning the multi-task model significantly outperformed the ablated model with the one-tailed version of the permutation test.

## Results

First, we evaluated the performance of the two pretrained models for their respective predictive targets. In the validation set, SFCN-multi achieved accurate age predictions (MAE=2.23, Figure 2a) but was slightly outperformed by SFCN-reg (MAE=2.21, Figure 2b), the latter tailored solely to predict brain age.

**Figure 2.**
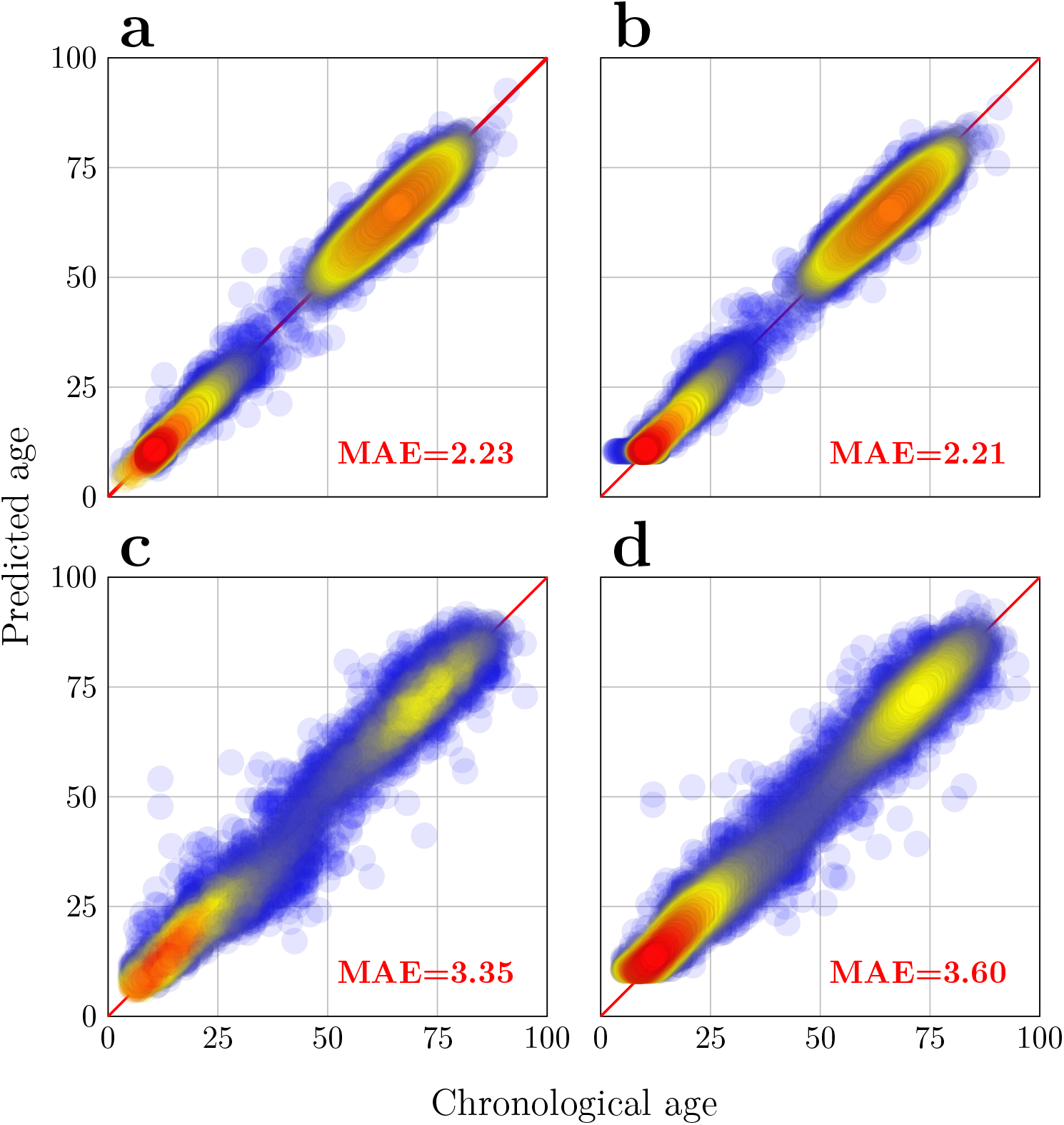
Predictive performance of the two pretrained models for predicting age. Performance was measured using the mean absolute error (MAE). Marker colour indicates density. (a) Performance of SFCN-multi in the validation set, containing data from the same sources that were used for training. (b) Performance of SFCN-reg in the validation set. (c) Performance of SFCN-multi in the test set, containing data from previously unseen sources. (d) Performance of SFCN-reg in the test set.

SFCN-multi also achieved accurate predictions for sex (AUC=0.99), but more moderate performance for the remaining targets, with an AUC of 0.61 for handedness, an MAE of 2.38 for BMI (baseline MAE=5.6), an R of 0.27 for fluid intelligence, and an R of 0.17 for neuroticism. In the held-out test data, SFCN-multi once again achieved accurate brain age predictions (MAE=3.35, Figure 2c), substantially outperforming SFCN-reg (MAE=3.6, Figure 2d). Here, SFCN-multi also achieved accurate out-of-sample predictions for sex (AUC=0.98), but only chance-level performance for handedness (AUC=0.48). For the remaining targets we did not retrieve a sufficient number of valid entries in the held-out test data to robustly measure performance.

### Multi-task pretraining results in a diverse feature space

To investigate the nature of the visual patterns the two models learned to recognise during pretraining, we extracted feature vectors from the bottleneck layer of both SFCN-reg and SFCN-multi for all participants in the test set. Assessing how the various entries in these vectors varied across participants, we observed that the feature space encoded by SFCN-reg contained highly intercorrelated features (mean absolute R≈0.92, Figure 3a), suggesting that all artificial neurons in this layer of the model were sensitive to highly covarying visual patterns, presumably encoding a relatively singular or global signal across the brain. For the features encoded by SFCN-multi the correlation was substantially lower (mean absolute R≈0.23, Figure 3b), a difference that persisted when residualising for age and sex (mean absolute R≈0.62 for SFCN-reg vs 0.17 for SFCN-multi, Supplementary Figure 6). Overall, these findings indicate that the latter model had learned to detect and leverage a broader range of morphological patterns.

**Figure 3.**
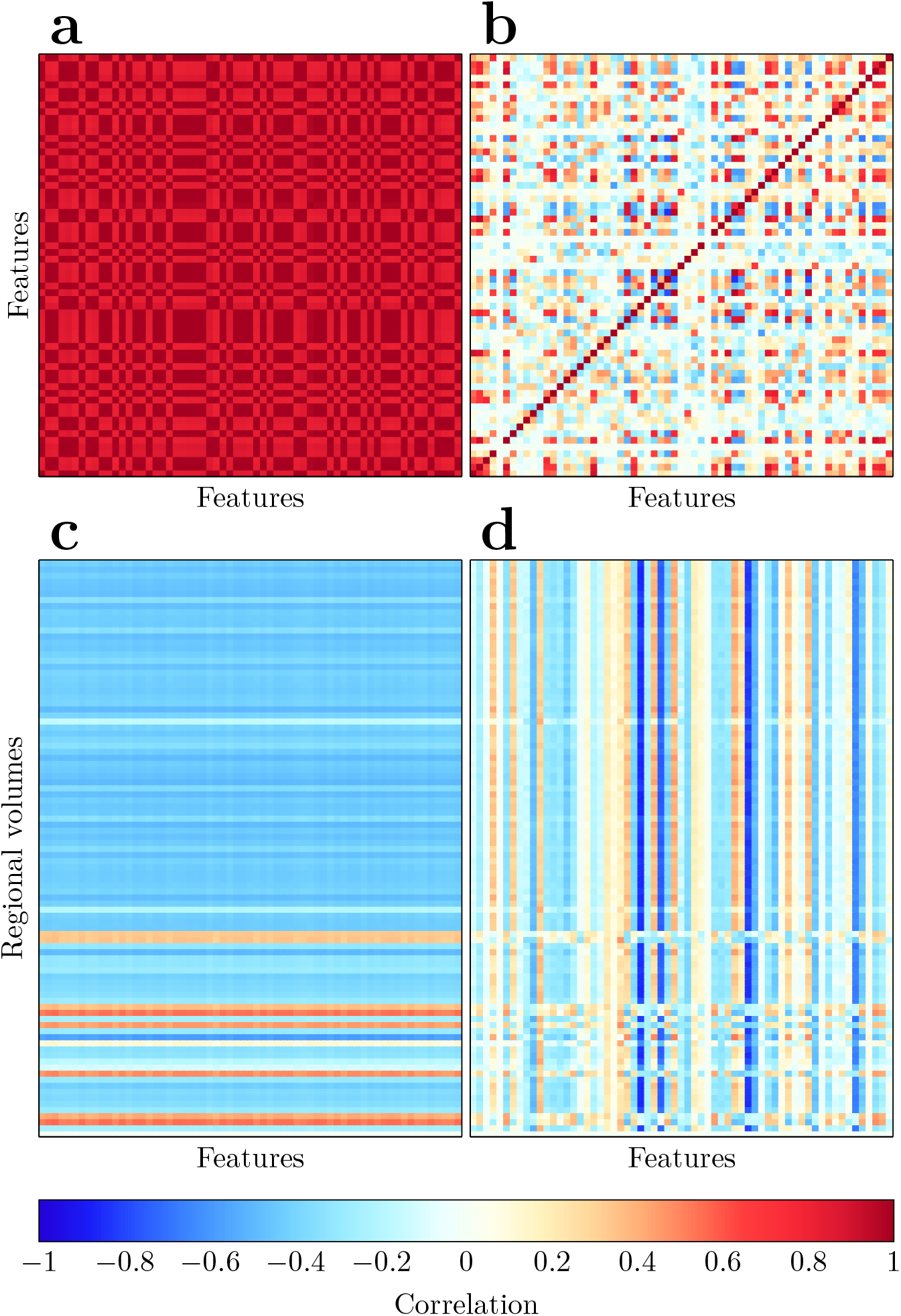
Visualisation of the feature spaces learned by SFCN-reg and SFCN-multi. (a) Intercorrelations between features from the second-to-last layer of SFCN-reg, calculated across all participants in the held-out test set. (b) Intercorrelations between bottleneck features from SFCN-multi. (c) Correlations between features from SFCN-reg and regional volumes calculated using FastSurfer in the same sample. (d) Correlations between features from SFCN-multi and regional volumes.

This impression was further corroborated when we investigated the correlations between the feature vectors from the two models and regional volumes derived from FastSurfer (Figure 3c and d). Features from SFCN-reg showed a high degree of stability per individual region across features, indicating that they generally reflected the same global volumetric signature. In contrast, correlations with features from SFCN-multi were stable across regions within each feature, indicating that individual features were broadly associated with volumetric differences with consistent polarity and magnitude. However, this also implies that the features covaried with different global volumetric patterns, emphasising their diversity. The apparent variability among the SFCN-multi correlations was quantified in a mean cosine distance between feature-wise correlation vectors of 0.85, as opposed to 0.002 for SFCN-reg, underscoring the differences between the feature spaces.

### Pretrained models yield superior predictive performance in a known downstream task

When training and applying new models to predict age in the heterogeneous dataset originating from a variety of unknown sources, both models based on the pretrained SFCN-multi and baseline models trained from scratch saw better performance with more training data (Figure 4a and Supplementary Figure 2). Largest absolute improvement from bigger samples, but also the worst overall performance, was achieved by the baseline models, with median RAEs decreasing monotonically from 0.63 (*n* = 100) to 0.23 (*n* = 2500). The finetuned models yielded second-to-worst performance, with median RAEs spanning from 0.20 (*n* = 100) to 0.15 (*n* = 2500). Best performance was achieved by the frozen and transferred models with median RAEs from 0.15 (*n* = 100) to 0.14 (*n* = 2500), the two being practically and statistically indistinguishable across all sample sizes. Overall, all three strategies relying on the pretrained SFCN-multi substantially and reliably outperformed the baseline models (*p* < 0.05 for all comparisons), and consistently required less epochs to converge (Figure 4b).

**Figure 4.**
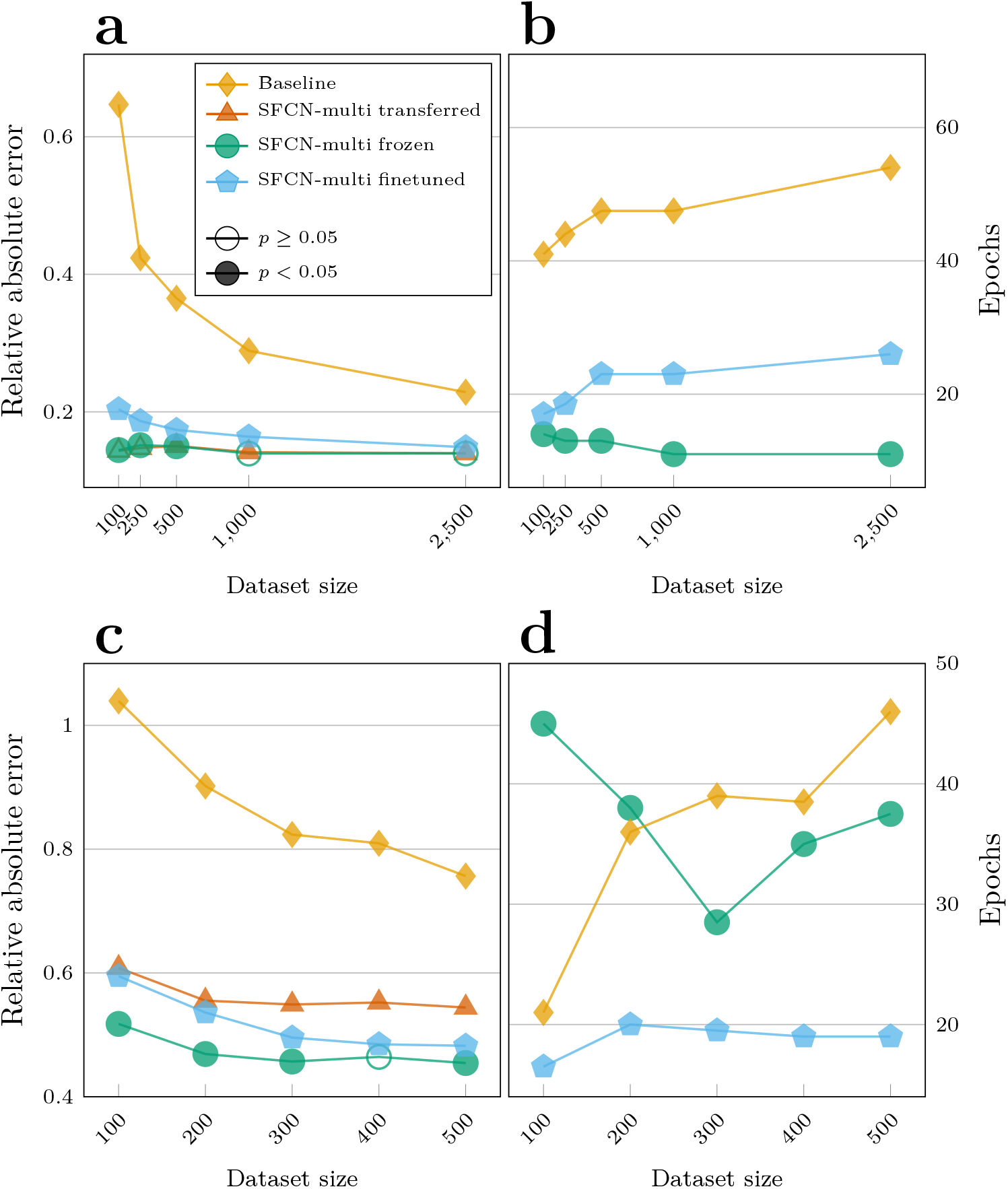
Performance of the models trained to predict age in previously unseen datasets. Each marker corresponds to the median across 50 bootstrapped runs per sample size and modelling strategy. (a) Predictive performance of the models in the heterogeneous dataset. Filled markers indicate that the given strategy achieved a statistically significant improvement (p<0.05 in a paired permutation test across the 50 runs) over the immediately lower-performing strategy at the given sample size. Note that the transferred and frozen models almost perfectly overlap (b) Number of epochs required for convergence in the hetero-geneous dataset. The transferred model did not require further training, and is not displayed. (c) Predictive performance of the models in the homogeneous dataset. (d) Number of epochs required for convergence in the homogeneous dataset.

Training models to predict age in the dataset constrained to be homogeneous in terms of scanning sites and age ranges, the overall pattern of performance was similar, with the pretrained models consistently outperforming the baseline models across all sample sizes (*p* < 0.05 for all comparisons, Figure 4c and Supplementary Figure 3). However, the relative difference between the two over-arching strategies was smaller, with the baseline models achieving median RAEs from 1.04 (*n* = 100) to 0.76 (*n* = 500), and the pretrained models spanning 0.61 (transferred models, *n* = 100) to 0.45 (frozen models, *n* = 500). Furthermore, the internal ranking among the pretrained models differed. Notably, the two strategies involving retraining, namely the frozen and finetuned models, consistently outperformed the directly transferred models, with the latter achieving median RAEs from 0.61 to 0.54. The frozen models performed best, with median RAEs from 0.52 (*n* = 100) to 0.45 (*n* = 500). Another difference in this dataset was that the finetuned models required substantially less training epochs than both the frozen and baseline models, and that the most efficient choice among the two latter relied on sample size (Figure 4d).

### Pretrained models yields superior predictive performance in new downstream tasks

To evaluate the efficacy of the pretraining and transfer learning schemes in novel downstream tasks, we first trained and evaluated models for AD vs HC classification. Here, the results were more mixed than for the age tasks (Figure 5a and Supplementary Figure 4), but some high level trends emerged. First, among the various deep learning strategies, finetuning SFCN-multi resulted in the best performance with median AUCs spanning 0.87 (*n* = 100) to 0.95 (*n* = 500), significantly outperforming the rest across all sample sizes (*p <* 0.05 for all comparisons). Marginally, but consistently, worse performance was achieved by finetuning SFCN-reg, with median AUCs spanning 0.86 to 0.94. Among the baseline and frozen models, the results were statistically indistinguishable for *n* ∈ {100, 200}, but the former outperformed the latter for *n* ≥ 300. Notably, the gap between the finetuned and baseline models decreased with larger sample sizes, although their ordering remained consistent. Interestingly, for *n* = 100, the linear models achieved a median AUC of 0.94, outperforming all CNNs (*p <* 0.05 for all comparisons). For *n* = 200 they also seemingly outperformed the deep learning alternatives with a median AUC of 0.92, however the relative improvement over finetuning SFCN-multi did not reach statistical significance. For *n* ≥ 300 they were significantly worse than the best CNNs. For this task, retraining SFCN-reg required substantially more compute than the remaining alternatives, while pretraining based on SFCN-multi were the most efficient strategies.

**Figure 5.**
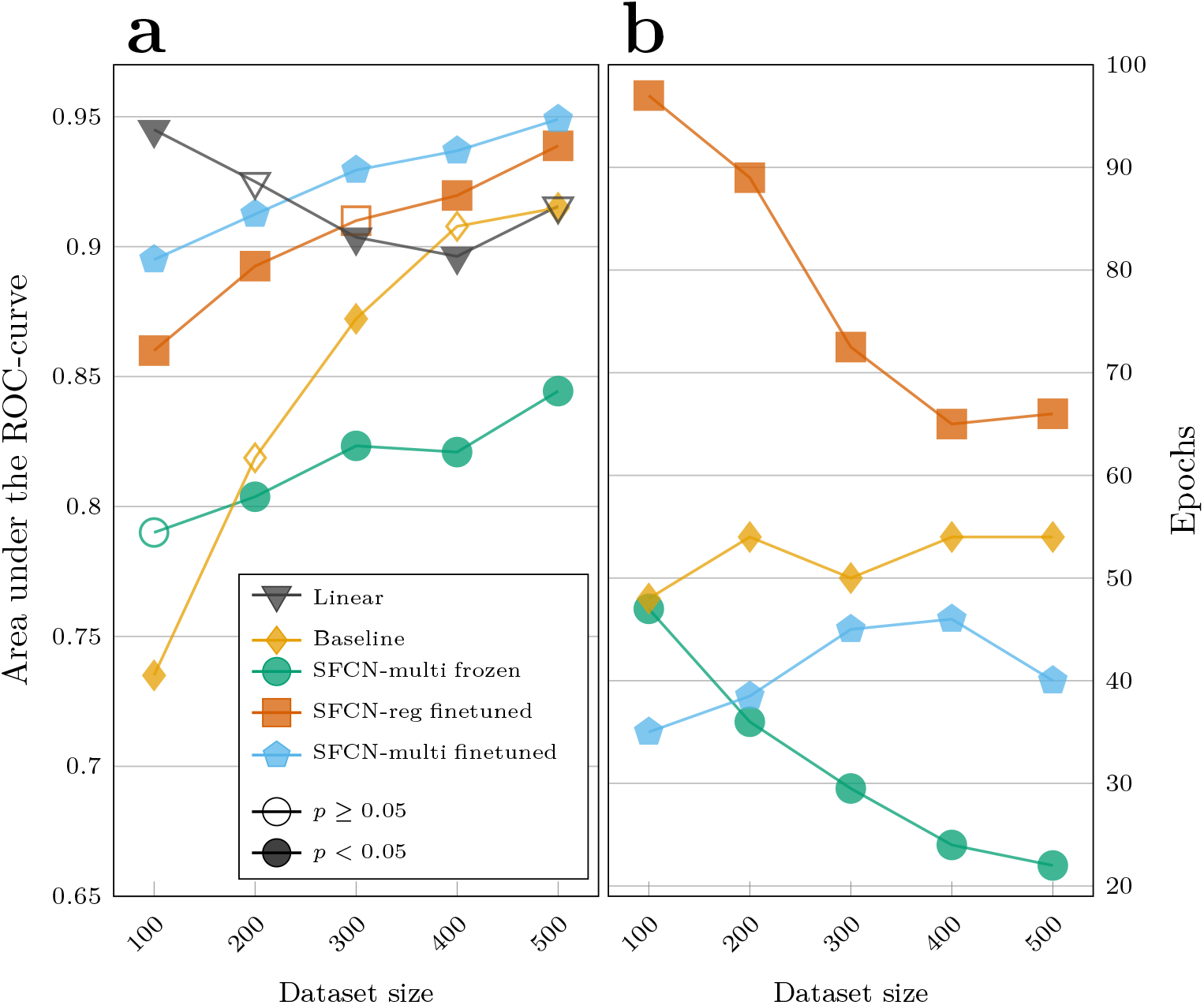
Performance of the various models trained to differentiate patients with Alzheimer’s disease and healthy controls. Each marker corresponds to the median across 50 bootstrapped runs per sample size and modelling strategy. (a) Predictive performance of the strategies. Filled markers indicate that the given strategy achieved a statistically significant improvement (p<0.05 in a paired permutation test across the 50 runs) over the immediately lower-performing strategy at the given sample size. (b) Number of epochs required for convergence.

Next, we evaluated the utility of our pretrained models to differentially diagnose SCZ and BD. Here, the overall performance of all models was significantly worse, with no strategy yielding a median AUC that surpassed 0.7 (Figure 6 and Supplementary Figure 5). Among the deep learning models, the finetuning strategy generally yielded the best performance with median AUCs spanning 0.61 (*n* = 100) to 0.68 (*n* = 500), except for *n* = 100 where the baseline models somewhat surprisingly outperformed them (median AUC=0.62). However, this performance difference was not significant (*p* > 0.05). For the remaining sample sizes, the finetuned models consistently outperformed the two other CNN alternatives (all *p* < 0.05). The linear models saw the overall largest performance gain in median AUC as the sample size increased, from a statistical chance-level of 0.5 for *n* = 100 to 0.69 for *n* = 400. Interestingly, the performance of these relatively simple models was once again on par, and appearingly even marginally better, than the finetuned models, representing the best deep learning alternative, for *n* ≥ 300. However, their relative improvement did not reach statistical significance (*p* > 0.05). Also in this task, the baseline CNNs consistently required more epochs to reach convergence than the two strategies relying on retraining SFCN-multi.

**Figure 6.**
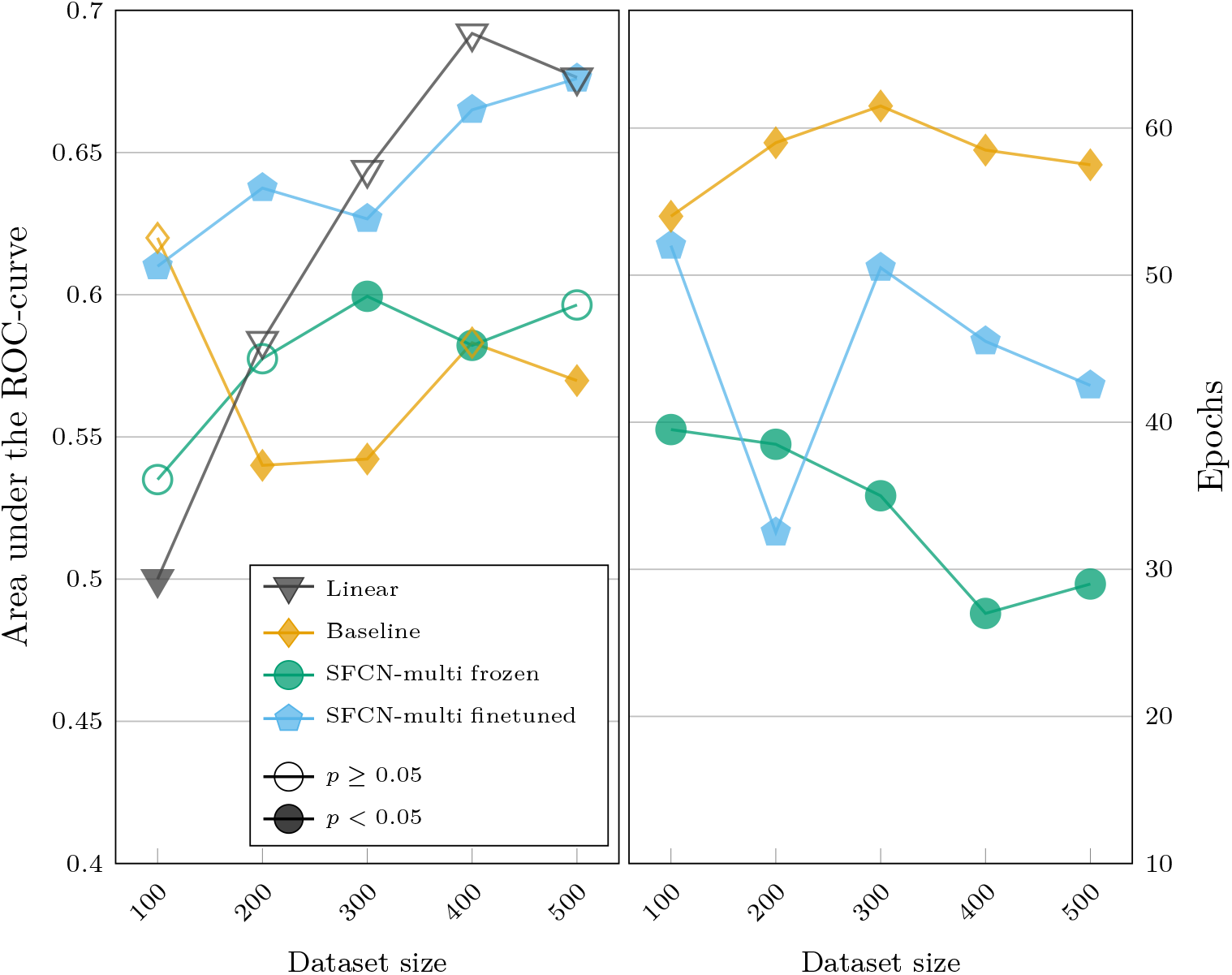
Performance of the various models trained to differentiate patients with schizophrenia and bipolar disorder. Each marker corresponds to the median across 50 bootstrapped runs per sample size and modelling strategy. (a) Predictive performance of the models. Filled markers indicate that the given strategy achieved a statistically significant improvement (p<0.05 in a paired permutation test across the 50 runs) over the immediately lower-performing strategy at the given sample size. (b) Number of epochs required for convergence.

### The importance of age as a pretraining target varies across downstream tasks

Lastly, we assessed the performance of the ablated models that were pretrained without age as a pretraining target. The model that simply had age removed did not see convergence for several of the remaining targets in the pretraining scheme, and was left out of the remaining analyses (see sub-panels on sex, handedness and BMI in Supplementary Figure 7a). The model that was pretrained with age scrambled saw no convergence for age, as expected, slightly worse predictive performance than the original multi-task model for sex and handedness, and better results for BMI, fluid intelligence and neuroticism (Supplementary Figure 7b) in the validation set.

In the AD vs HC classification task, transfer learning based on the model pretrained with scrambled age achieved a median AUC of 0.84 (Supplementary Figure 8a), significantly lower (all p<0.05) than both the baseline model (AUC=0.87), and the two competing transfer learning approaches relying on finetuning the brain age model (AUC=0.91) and the ordinary multi-task model (AUC=0.93). For the SCZ vs BD classification task, finetuning the pretrained model with scrambled age achieved an AUC of 0.65 (Supplementary Figure 8b), significantly outperforming both the baseline (AUC=0.54, p<0.05 in a one-sided test) and finetuned brain age (AUC=0.55, p<0.05 in a two-sided test) models, and performing on par with the finetuned multi-task models (AUC=0.63, p>0.05 in a one-sided test).

## Discussion

In this work, we pretrained a multi-task CNN using a state-of-the-art architecture on a large dataset of T1-weighted structural MRIs. We showed that our multi-task pretraining scheme produced a model with a richer feature space than an equivalent CNN pretrained solely to predict age, and that this diversity yielded more accurate age predictions in held-out datasets. We also exhibited how transfer learning from models pretrained in large samples outperformed models trained from scratch in downstream predictive tasks, both when the task was incorporated in the pretraining scheme, and for new classification problems. In sum, our work demonstrates the usefulness of models pretrained in large neuroimaging datasets, and shows that multi-task learning is an attractive option to enhance their generalisability, addressing limitations demonstrated in common single-task alternatives (Tan et al., 2025).

Where previous studies have suggested the general efficacy of pretraining CNNs for neuroimaging data (Agarwal et al., 2021; Ardalan & Subbian, 2022; Valverde et al., 2021), or shown that pretrained models outperform randomly intialised ones in specific use-cases (Bashyam et al., 2020; Dufumier et al., 2024), our work is among the most comprehensive inquiries on their utility to date. And while the downstream tasks examined here represent a small subset of the possible applications of CNNs to neuroimaging data, our results show some clear trends, paving way for best practices. First and foremost, transfer learning outperformed models trained from scratch for all downstream tasks and sample sizes, except a single instance where the two approaches yielded results that were statistically indistinguishable. This emphasizes the benefits of starting with a pretrained model when working with small datasets, when such a model exists. However, as others have reported the possibility of detrimental effects of transfer learning (Z. Wang et al., 2019), it is worth underscoring that this relies on some level of similarity between the pretraining and downstream task. We saw supportive evidence of this in our ablation study, where the negative transfer effects of pretraining with scrambled age labels yielded worse results than training models from scratch when classifying patients with AD and HCs. Furthermore, echoing prior findings (Valverde et al., 2021), we observed that the performance advantage of transfer learning diminished with increasing sample sizes, suggesting the existence of an upper bound where pretraining no longer is beneficial.

In addition to comparing various transfer learning strategies against randomly initialised models, we benchmarked the accuracy of off-the-shelf brain age predictions from our pretrained multi-task model against various models trained to predict age in new datasets, tested in those same datasets. Here, the optimal strategy seemed to require some level of dataset-specific retraining. However, the off-the-shelf model performed comparatively to the transfer learning approaches and substantially outperformed the models trained from scratch. This implies that when a pretrained model trained to predict the target of interest in a sufficiently large and heterogeneous dataset exists, using it as-is is often a reasonable approach. Furthermore, although the off-the-shelf predictions were somewhat less precise than those from the transfer learning approaches, group-level accuracy in itself is rarely the goal of brain age studies (Jirsaraie et al., 2023). Instead these studies often utilise the brain age gap (BAG), encoding the difference between apparent brain age and chronological age (Franke et al., 2010), as a proxy for brain health, and others have argued that the most informative BAGs originate from models that are not necessarily the most accurate (Bashyam et al., 2020; Schulz et al., 2024). It is tempting to speculate whether the brain age predictions from the multi-task model, constructed to reflect a broader set of underlying morphological patterns, encodes more interesting variability than those stemming from a finetuned single-task brain age model.

While the main focus of this work was to investigate the usefulness of pretraining CNNs on neuroimaging data, the behaviour of the linear models was striking. First, when assessed separately across the two tasks, their predictive behaviour differed markedly: for classifying AD patients vs HCs, performance appeared relatively stable as the amount of training data increased, whereas for differentiating SCZ from BD patients the results almost monotonically improved as a function of sample size. Comparing their performance relative to the deep learning models complicates the picture further: for AD vs HC, linear models outperformed CNNs with the least amount of training data, but were eventually overtaken as the datasets grew in size. For SCZ vs BD this was reversed, with pretrained CNNs performing the best in the smallest samples, but being overtaken the linear models with more training data.

We can only speculate about the underlying reasons for these findings, but it is intriguing to contemplate whether they reflect characteristics related to the populations and ontologies comprising the two classification problems. For AD vs HC, the stable performance of the linear models suggests that there are relatively simple volumetric differences that allow for a reasonably accurate linear separation of the two groups. This is corroborated by earlier reports of minimalistic models achieveing noteworthy predictive performances for differentiating these two groups (Grødem et al., 2024), and the etiological understanding of AD (Pini et al., 2016). The relative performance of CNNs at this task could reflect their propensity to underperform in low signal-to-noise regimes (Schulz et al., 2020), particularly in small datasets. However, with sufficient amounts of data they were able to leverage non-linear characteristics inaccessible to the linear models, yielding more accurate predictions. For SCZ vs BD, the positive trend in predictive performance of the linear models suggest that there are volumetric differences that allow for some level of differentiation between these patient cohorts, but that these are less prominent. Furthermore, it is worth emphasizing that the best models for this task, i.e. the linear models trained with the largest sample sizes, did not reach AUCs surpassing 0.7. Both of these findings support the prevalent notion that these are highly heterogeneous and complex conditions (Alnæs et al., 2019; Wolfers et al., 2018) with overlapping morphological signatures (Doan et al., 2017; Schwarz et al., 2019). The fact that pretrained CNNs robustly outperformed linear models with the smallest amount of training data could speak to the efficacy of pretraining, that the non-linear condensation of morphological variability learned by the models in population data is beneficial also for differentiating the two groups. However, the linear models eventually matching their predictive performance indicate that this initial advantage is at some point outweighed by the intricacy of manouvering heterogeneity with presumably low signal-to-noise ratios. Concluding decisively whether the different performance patterns of these remarkably different model types relay some ontological insights will require further, more targeted, investigation.

Previous studies have challenged the notion that deep learning models should be the universally preferred choice for analysing neuroimaging data (Dufumier et al., 2024; T. He et al., 2020; Schulz et al., 2020). Our results contribute to the empirical basis underlying this debate in two ways. First, by supporting the idea that CNNs do not trivially represent a free lunch for predicting clinical phenotypes from structural MRIs, being outperformed by linear models in several direct comparisons. Determining the optimal model appears contingent on a combination of sample size, heterogeneity, and signal-to-noise ratio. However, our results also corroborate that pretraining seems to result in an overall performance improvement for CNNs (Dufumier et al., 2024), effectively nudging the scale in their direction. This underscores the promise of combining highly expressive models with large heterogeneous datasets comprised of more or less healthy participants, to uncover patterns that have predictive value in clinical cohorts. Overall, this represents an encouraging perspective on how deep learning can contribute to advancing clinical neuroscience as a whole.

Accompanying the positive outlook above are several limitations to the present study that should be considered. First, in terms of the construction of the pretrained models, and the apparent efficacy of the multi-task learning scheme. We refer to our two pretraining alternatives as a multi-task model and a single-task brain age model, implying that their main difference lies in their respective predictive targets. Here, it is worth noting that there are also architectural differences between them, i.e. the choice of global pooling strategy and the application of dropout during training. These are both known to contribute to feature diversity, motivating their inclusion here, and we cannot disentangle their partial effects from the multi-task target in itself. Relatedly, we employed a single hyperparameter setting for each model type during pretraining, and a broader search could yield different results. And finally, we selected a relatively simple preprocessing pipeline to make the model as accessible as possible, although previous findings indicate that some preprocessing can be beneficial, also when using deep learning (Dufumier et al., 2024). Nonetheless, we believe the impact of these design choices on our overall conclusions to be negligible. Continuing, although we devised our downstream tasks to be somewhat representative, the extent of their spread was limited by practicalities. It is not trivially true that our findings generalise to other, more deviant, tasks, or in substantially different populations. The need to run thousands of models in a relatively unified way also severely limited the opportunity to make precise decisions about individual modelling tasks. While we believe our relatively crude hyperparameter search performed individually for each bootstrapped sample for each downstream task is a reasonable compromise, we believe that putting more effort into each individual model would have lifted the overall predictive performance across the board. Nonetheless, we see no principled reason this would affect the comparison between the high-level approaches examined here in a biased manner, except maybe underestimating the general efficacy of deep learning models that are more configurable than their linear counterparts. Finally, our pretraining approach was shaped by pragmatism, recognisable in our choice of weighting scheme, the choice of multi-task targets, and the effort put into creating datasets that were reasonably, though not exhaustively, complete in terms of variable coverage. Since the primary objective of this work was to examine the potential of multi-task pretraining, we believe that these choices minimally affect our general conclusions regarding downstream efficacy. They do, however, substantially influence the relative predictive performance across pretraining targets. Most notably, due to our selection process prioritizing variables based on participant-wise coverage, several of the pretraining targets were almost exclusively retrieved from the largest datasets, i.e. UKBB and ABCD. Therefore, this specific aspect of our results should be interpreted with the utmost caution.

To conclude, we compiled a large dataset of T1-weighted structural MRIs to pretrain a multi-task CNN and demonstrated its usefulness, paving the way for more efficient and accurate modelling based on neuroimaging data through transfer learning. The pretrained model and interfaces for its use is available on GitHub, HuggingFace, and DockerHub.

## Data Availability

The data incorporated in this study was compiled from a variety of sources. Requests for the underlying data need to be placed with the principal investigators for each individual study.

## Acknowledgements

The data used in this study was compiled from several sources, each with their own acknowledgements that can be found in Supplementary Table 1. The work was funded by the Norwegian Research Council (grant numbers 324499). All analyses were performed on the Services for Sensitive Data (TSD, project p33), University of Oslo, Norway, with resources from UNINETT Sigma2–the National Infrastructure for High-Performance Computing and Data Storage in Norway.

## Data availability

The data incorporated in this study was compiled from a variety of sources. Requests for the underlying data need to be placed with the principal investigators for each individual study. An overview of these data sources and their origins can be found in Supplementary Table 1.

## Code availability

The code and weights for the pretrained model and various interfaces for interacting with it is available on https://github.com/estenhl/pyment-public, https://huggingface.co/estenhl/pyment-public, and https://hub.docker.com/u/estenhl.

