## Supplementary Figures 1-8 and Tables 1-5 for "Learning diverse and generic representations of the brain with large-scale multi-task pretraining"

Leonardsen et. al, 2025

5 Correlation between the targets used for multi-task pretraining . 26

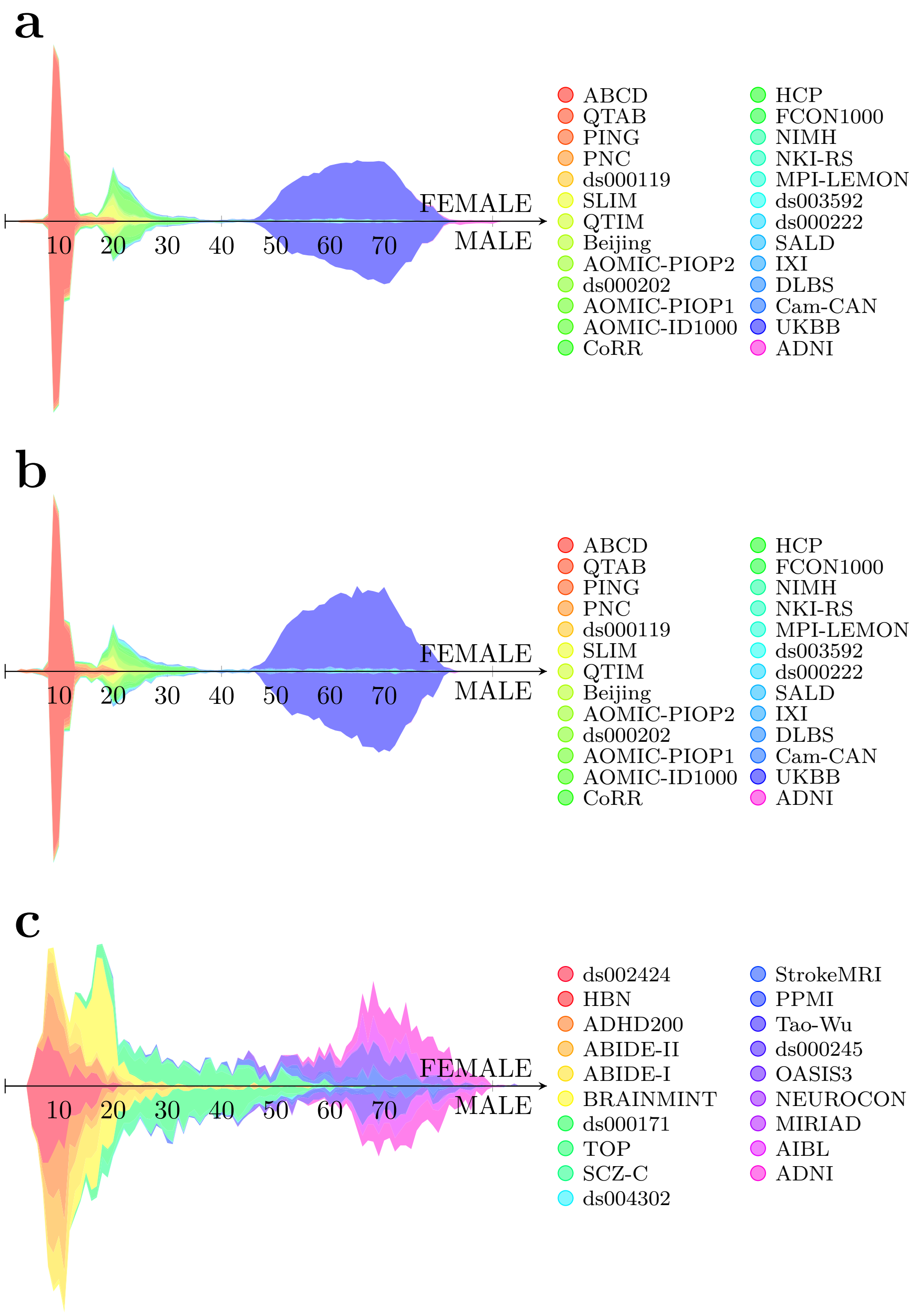

Supplementary Figure 1: Age and sex distributions in the datasets used for (a) training, (b) validating and (c) testing the pretrained models.

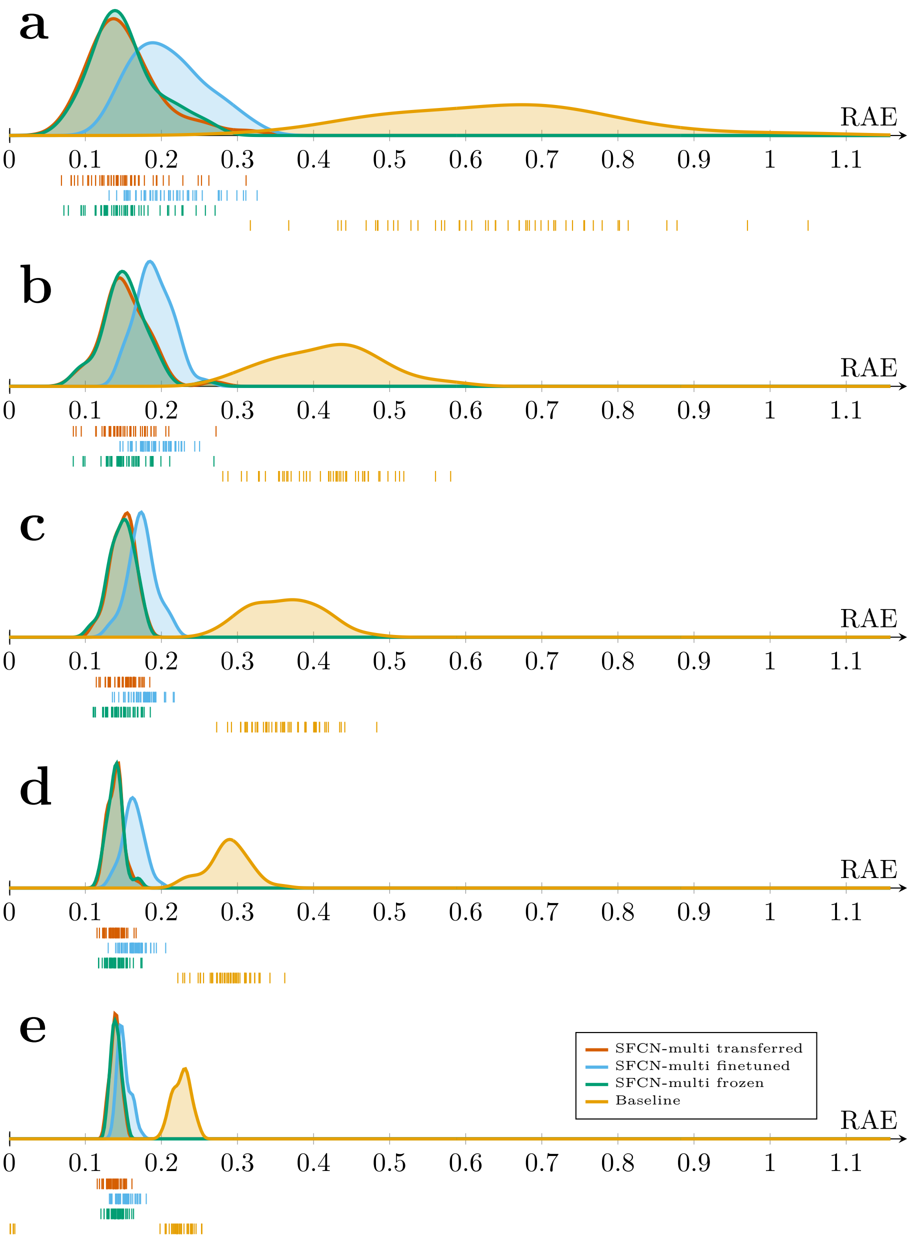

Supplementary Figure 2: Distributions of the out-of-sample Relative Absolute Errors (RAEs) for the models trained to predict age in the heterogeneous dataset, split by strategy. The models were trained on bootstrapped samples of size (a) $n = 100$, (b) $n = 250$, (c) $n = 500$, (d) $n = 1000$, and (e) $n = 2500$. The points below the x-axis represents RAEs achieved by individual models of the given type.

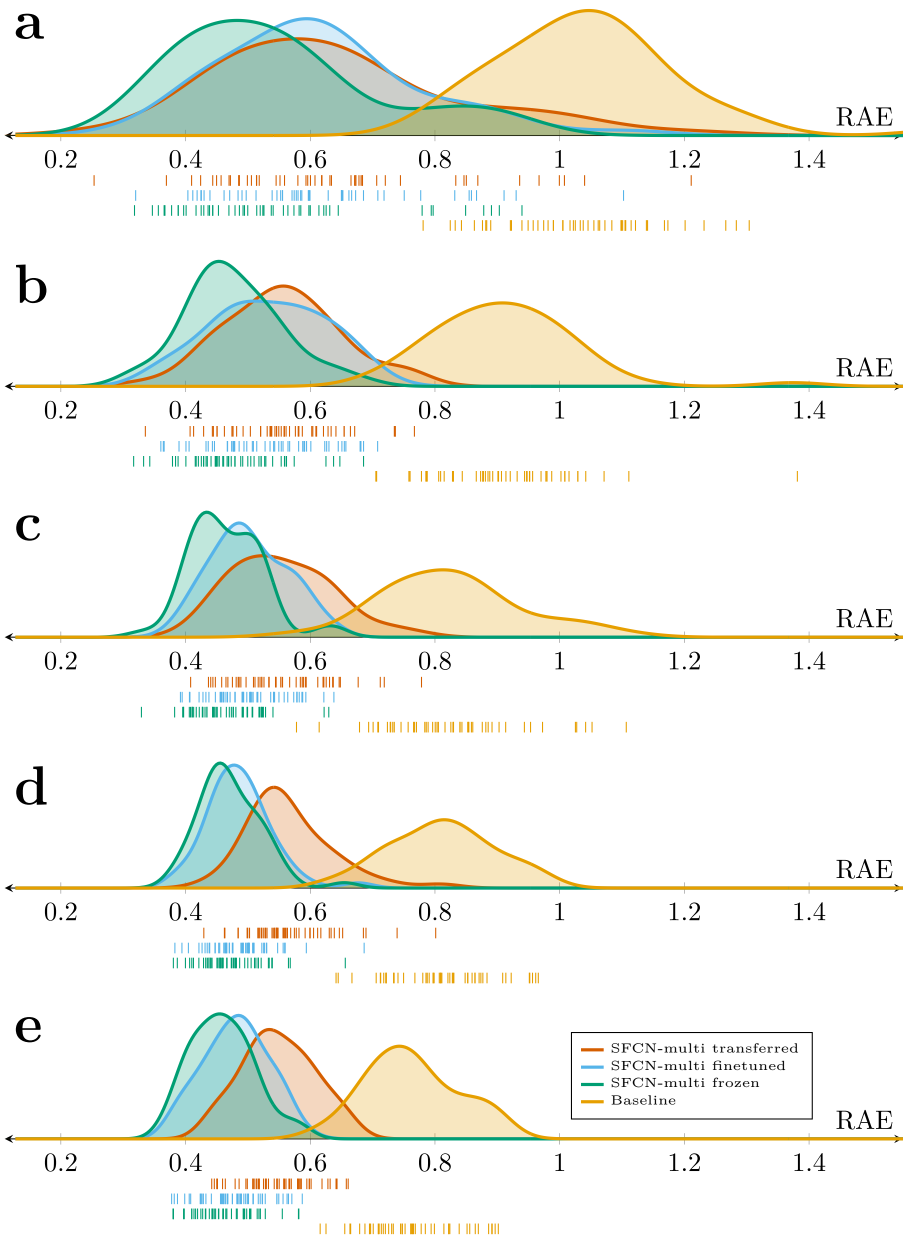

Supplementary Figure 3: Distributions of the out-of-sample Relative Absolute Errors (RAEs) for the models trained to predict age in the homogeneous dataset, split by strategy. The models were trained on bootstrapped samples of size (a) $n = 100$, (b) $n = 200$, (c) $n = 300$, (d) $n = 400$, and (e) $n = 500$. The points below the x-axis represents RAEs achieved by individual models of the given type.

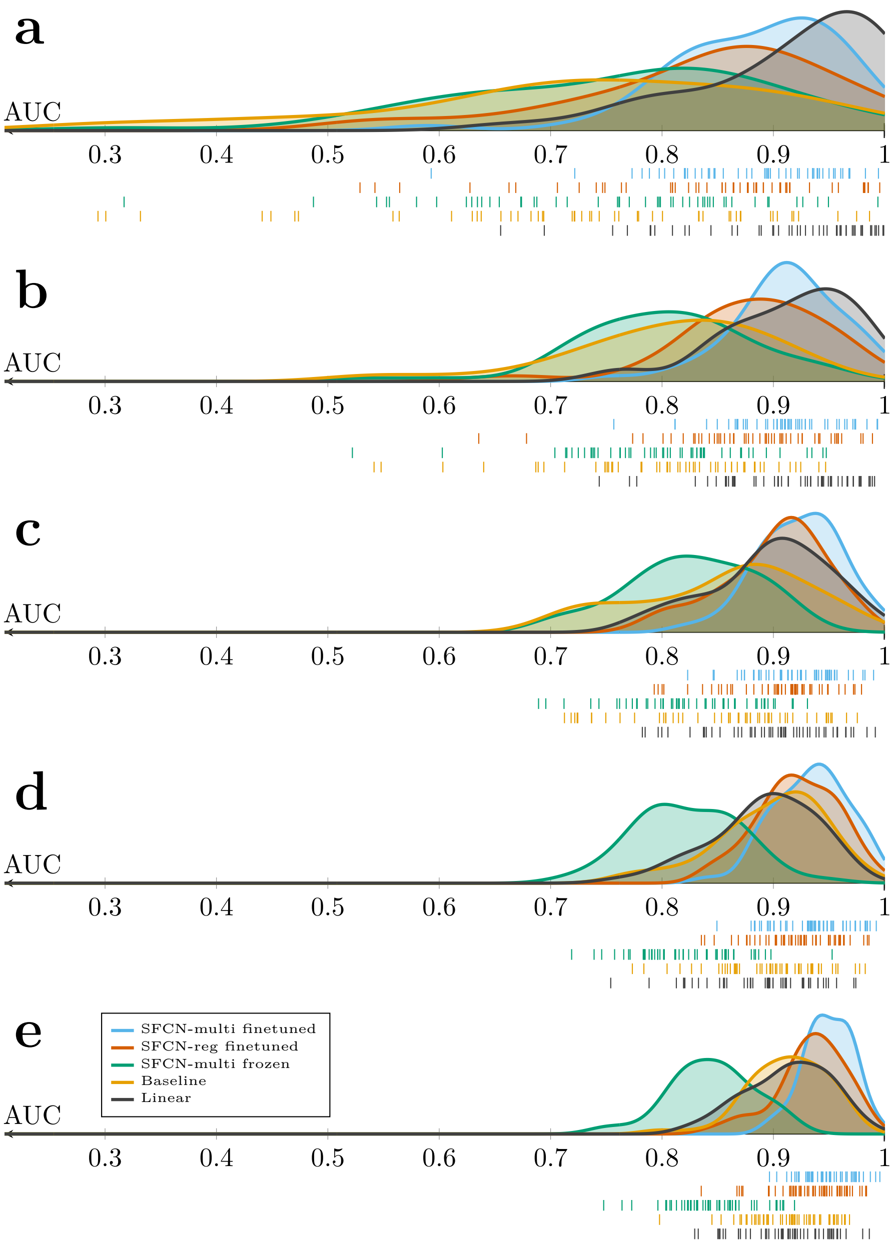

Supplementary Figure 4: Distributions of the out-of-sample areas under the ROC curve (AUCs) for the six different models trained to differentiate patients with Alzheimer's disease and healthy controls, split by strategy. The models were trained on bootstrapped samples of size (a) $n = 100$, (b) $n = 200$, (c) $n = 300$, (d) $n = 400$, and (e) $n = 500$. The points below the x-axis represents AUCs achieved by individual models of the given type.

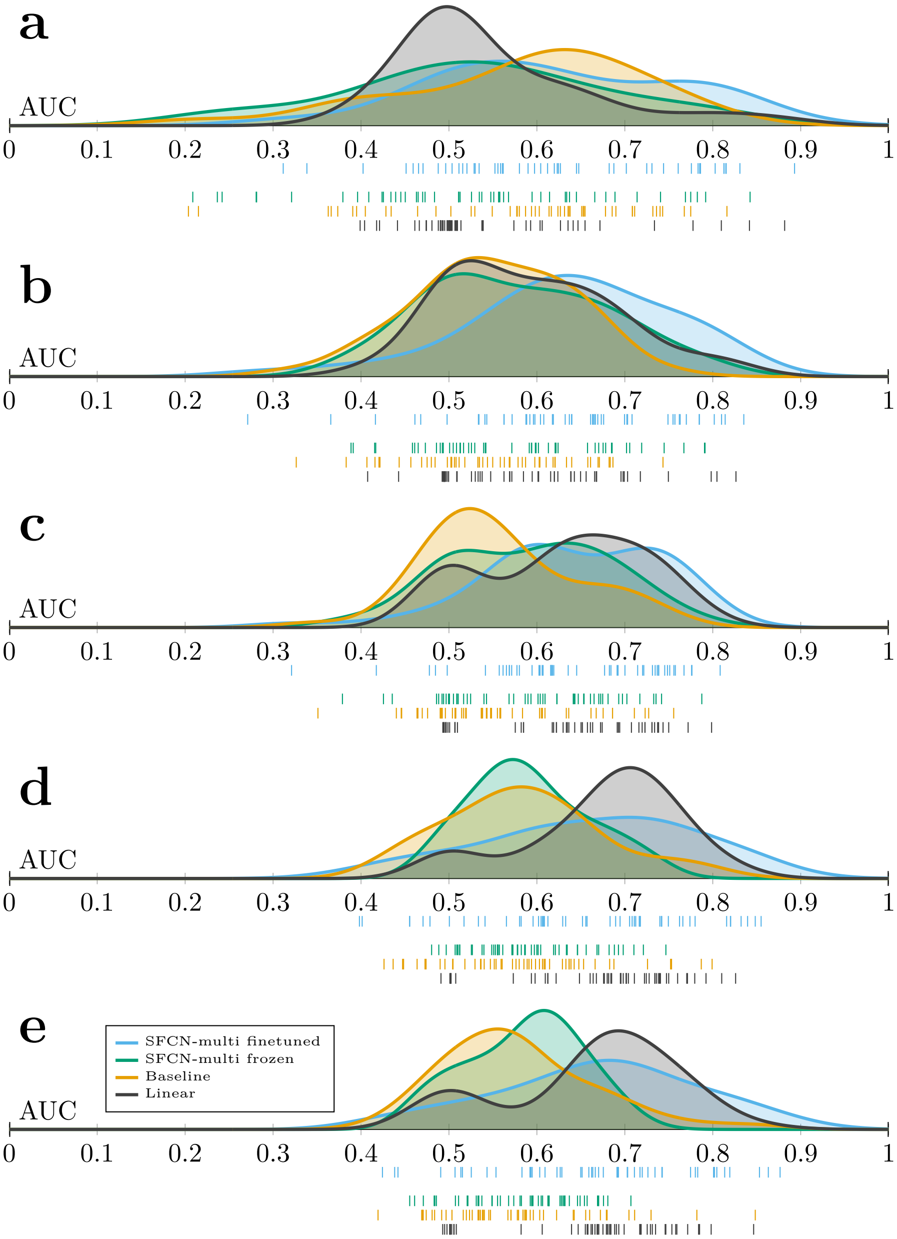

Supplementary Figure 5: Distributions of the out-of-sample areas under the ROC curve (AUCs) for the six different models trained to differentiate patients with schizophrenia and bipolar disorder, split by strategy. The models were trained on bootstrapped samples of size (a) $n = 100$, (b) $n = 200$, (c) $n = 300$, (d) $n = 400$, and (e) $n = 500$. The points below the x-axis represents AUCs achieved by individual models of the given type.

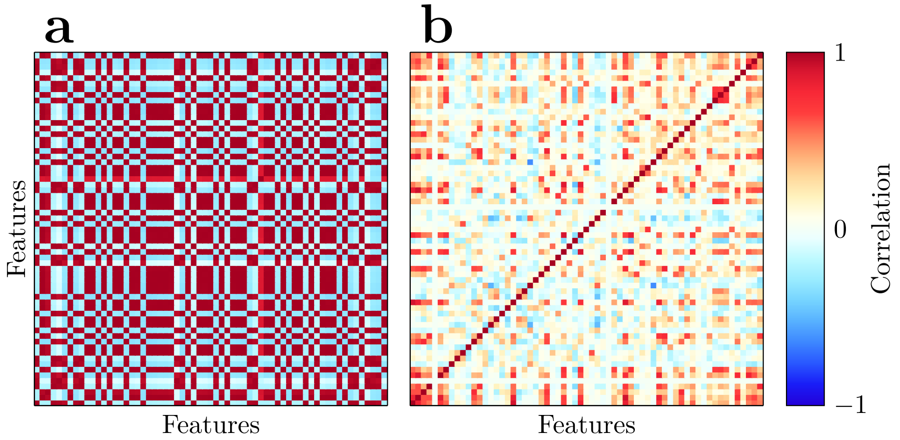

Supplementary Figure 6: Visualisation of the feature spaces learned by SFCN-reg and SFCN-multi, after residualising for age and sex. (a) Intercorrelations between features from the second-to-last layer of SFCN-reg, calculated across all participants in the held-out test set. (b) Intercorrelations between bottleneck features from SFCN-multi.

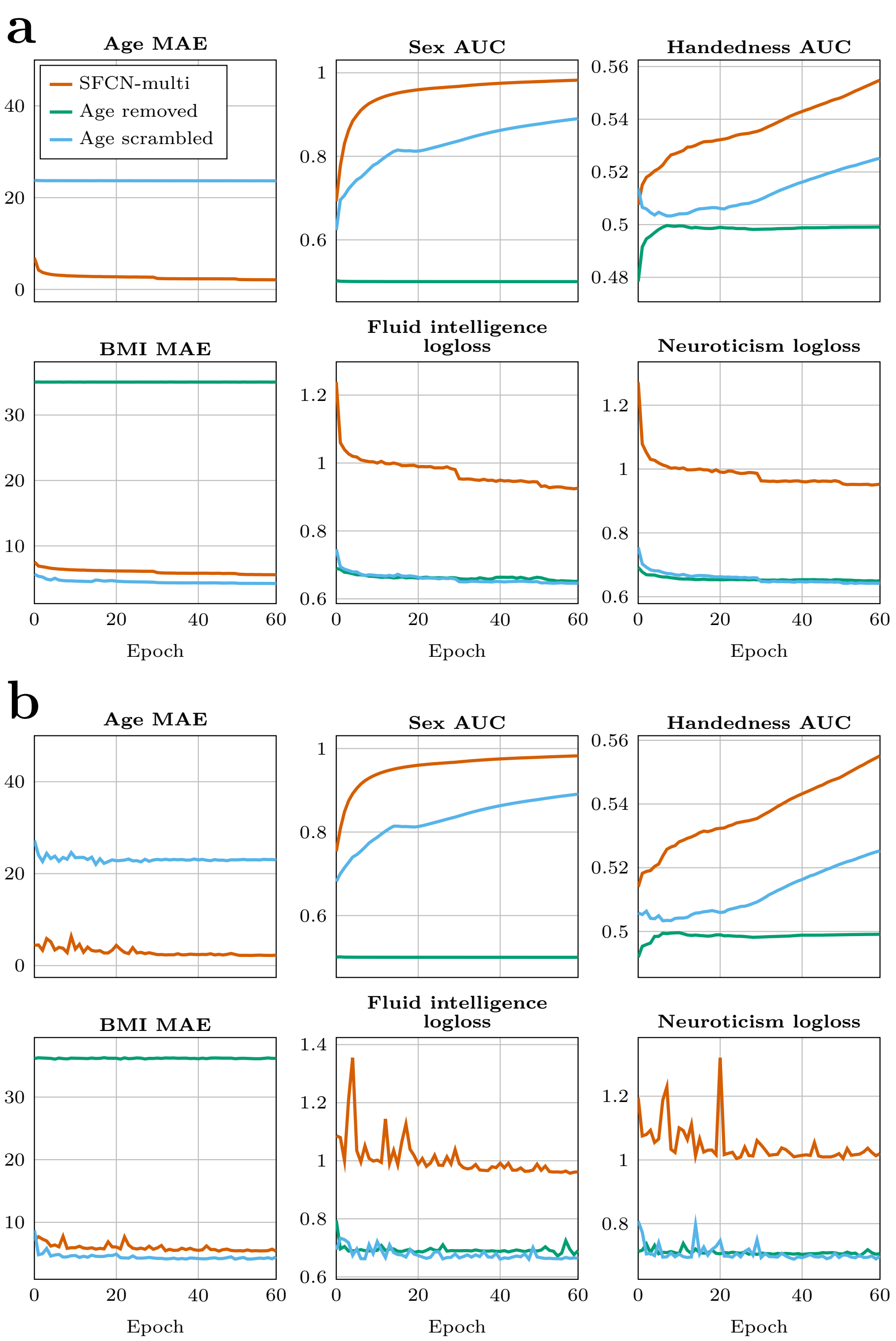

Supplementary Figure 7: (a) Training and (b) validation performance across the different multi-task targets for the three models in the ablation study on the importance of age for pretraining.

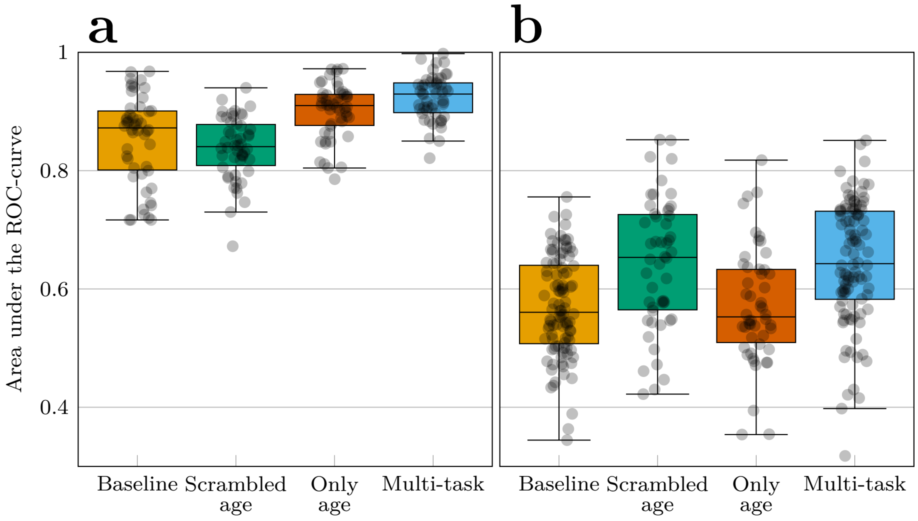

Supplementary Figure 8: Performance of the ablated pretrained models without age for (a) classifying patients with Alzheimer's disease and healthy controls and (b) classifying patients with schizophrenia and bipolar disorder. Each dot represents the performance of a single model.

| Name | Short-hand | Source | Acknowledgements | References |
|---|---|---|---|---|
| Adolescent Brain Cognitive Development | ABCD | https://abcdstudy.org/ | Data used in the preparation of this article were obtained from the Adolescent Brain Cognitive DevelopmentSM (ABCD) Study (https://abcdstudy.org), held in the NIMH Data Archive (NDA). This is a multisite, longitudinal study designed to recruit more than 10,000 children age 9-10 and follow them over 10 years into early adulthood. The ABCD Study® is supported by the National Institutes of Health and additional federal partners under award numbers U01DA041048, U01DA050989, U01DA051016, U01DA041022, U01DA051018, U01DA051037, U01DA050987, U01DA041174, U01DA041106, U01DA041117, U01DA041028, U01DA041134, U01DA050988, U01DA051039, U01DA041156, U01DA041025, U01DA041120, U01DA051038, U01DA041148, U01DA041093, U01DA041089, U24DA041123, U24DA041147. A full list of supporters is available at https://abcdstudy.org/federal-partners.html. A listing of participating sites and a complete listing of the study investigators can be found at https://abcdstudy.org/consortium_members/. ABCD consortium investigators designed and implemented the study and/or provided data but did not necessarily participate in the analysis or writing of this report. This manuscript reflects the views of the authors and may not reflect the opinions or views of the NIH or ABCD consortium investigators. Our access to ABCD research data is given by Data Use Certification 22719. | Casey et al., 2018 |
| Autism Brain Imaging Data Exchange I | ABIDE-I | https://fcon_1000.projects.nitrc.org/indi/abide/abide_I.html | Primary support for the work by Adriana Di Martino was provided by the NIMH (K23MH087770) and the Leon Levy Foundation. Primary support for the work by Michael P. Milham and the INDI team was provided by gifts from Joseph P. Healy and the Stavros Niarchos Foundation to the Child Mind Institute, as well as by an NIMH award to MPM (R03MH096321). | Di Martino et al., 2014 |
| Autism Brain Imaging Data Exchange II | ABIDE-II | https://fcon_1000.projects.nitrc.org/indi/abide/abide_II.html | Primary support for the work by Adriana Di Martino and her team was provided by the National Institute of Mental Health (NIMH 5R21MH107045). Primary support for the work by Michael P. Milham and his team provided by the National Institute of Mental Health (NIMH 5R21MH107045); Nathan S. Kline Institute of Psychiatric Research. Additional Support was provided by gifts from Joseph P. Healey, Phyllis Green and Randolph Cowen to the Child Mind Institute. | Di Martino et al., 2017 |
| ADHD-200 | ADHD200 | https://fcon_1000.projects.nitrc.org/indi/adhd200/ | F. Xavier Castellanos, David Kennedy, Michael Milham, and Stewart Mostofsky are | Brown et al., 2012; Milham et al., 2012 |

| | | | | |
|---|---|---|---|---|
| | | | responsible for the initial conception of the ADHD-200 Consortium. Consortium steering committee includes Jan Buitelaar, F. Xavier Castellanos, Dan Dickstein, Damien Fair, David Kennedy, Beatriz Luna, Michael Milham (Project Coordinator), Stewart Mostofsky, and Julie Schweitzer. Data aggregation and organization was coordinated by the INDI team, which included Saroja Bangaru, David Gutman, Maarten Mennes, and Michael Milham. Web infrastructure and data storage were coordinated by Robert Buccigrossi, Albert Crowley, Christian Hasselgrove, David Kennedy, Kimberly Pohland, and Nina Preuss. The ADHD-200 Global Competition Coordinators were Damien Fair (Chair of Selection Committee, Editor in Chief for Global Competition Special issue) and Michael Milham. | |
| Alzheimer's Disease Neuroimaging Initiative | ADNI | https://adni.loni.usc.edu/ | Data used in the preparation of this article were obtained from the Alzheimer's Disease Neuroimaging Initiative (ADNI) database (adni.loni.usc.edu). The ADNI was launched in 2003 as a public-private partnership, led by Principal Investigator Michael W. Weiner, MD. The original goal of ADNI was to test whether serial magnetic resonance imaging (MRI), positron emission tomography (PET), other biological markers, and clinical and neuropsychological assessment can be combined to measure the progression of mild cognitive impairment (MCI) and early Alzheimer's disease (AD). The current goals include validating biomarkers for clinical trials, improving the generalizability of ADNI data by increasing diversity in the participant cohort, and to provide data concerning the diagnosis and progression of Alzheimer's disease to the scientific community. For up-to-date information, see adni.loni.usc.edu. Data collection and sharing for the Alzheimer's Disease Neuroimaging Initiative (ADNI) is funded by the National Institute on Aging (National Institutes of Health Grant U19AG024904). The grantee organization is the Northern California Institute for Research and Education. In the past, ADNI has also received funding from the National Institute of Biomedical Imaging and Bioengineering, the Canadian Institutes of Health Research, and private sector contributions through the Foundation for the National Institutes of Health (FNIH) including generous contributions from the following: AbbVie, Alzheimer's Association; Alzheimer's Drug Discovery Foundation; Araclon Biotech; BioClinica, Inc.; Biogen; BristolMyers Squibb | Mueller et al., 2005 |

| | | | | |
|---|---|---|---|---|
| | | | Company; CereSpir, Inc.; Cogstate; Eisai Inc.; Elan Pharmaceuticals, Inc.; Eli Lilly and Company; EuroImmun; F. Hoffmann-La Roche Ltd and its affiliated company Genentech, Inc.; Fujirebio; GE Healthcare; IXICO Ltd.; Janssen Alzheimer Immunotherapy Research & Development, LLC.; Johnson & Johnson Pharmaceutical Research & Development LLC.; Lumosity; Lundbeck; Merck & Co., Inc.; Meso Scale Diagnostics, LLC.; NeuroRx Research; Neurotrack Technologies; Novartis Pharmaceuticals Corporation; Pfizer Inc.; Piramal Imaging; Servier; Takeda Pharmaceutical Company; and Transition Therapeutics. | |
| Australian Imaging Biomarkers and Lifestyle Study | AIBL | https://aibl.org.au/ | Data used in the preparation of this article was obtained from the Australian Imaging Biomarkers and Lifestyle flagship study of ageing (AIBL) funded by the Commonwealth Scientific and Industrial Research Organisation (CSIRO) which was made available at the ADNI database (www.loni.usc.edu/ADNI). The AIBL researchers contributed data but did not participate in analysis or writing of this report. AIBL researchers are listed at www.aibl.csiro.au. | Ellis et al., 2009 |
| Beijing Normal University - Enhanced Sample | Beijing | https://fcon_1000.projects.nitrc.org/indi/retro/BeijingEnhanced.html | Financial support for the data used in this project was provided by a grant from the National Natural Science Foundation of China: 30770594 and a grant from the National High Technology Program of China (863): 2008AA02Z405. | Tian et al., 2011; Yan and Zang, 2010 |
| Brain and minds in transition | BRAIN-MINT | Authors | Accessed with approval from the Regional Committee for Medical Research Ethics South East Norway (REC, application number: 2019/943). Funded by the European Union (ERC, BRAINMINT, 802998) | |
| Cambridge Centre for Ageing and Neuroscience dataset | Cam-CAN | https://cam-can.mrc-cbu.cam.ac.uk/dataset/ | Data used in the preparation of this work were obtained from the CamCAN repository (available at http://www.mrc-cbu.cam.ac.uk/datasets/camcan/). Data collection and sharing for this project was provided by the Cambridge Centre for Ageing and Neuroscience (CamCAN). CamCAN funding was provided by the UK Biotechnology and Biological Sciences Research Council (grant number BB/H008217/1), together with support from the UK Medical Research Council and University of Cambridge, UK. | Shafto et al., 2014; Taylor et al., 2017 |
| Consortium for Reliability and Reproducibility | CoRR | https://fcon_1000.projects.nitrc.org/indi/CoRR/html/index.html | The National Institute on Drug Abuse and the National Natural Science Foundation of China (NSFC) have been instrumental in the CoRR collaboration providing the necessary funding and manpower to build the foundation of the project along with the Child Mind Institute, the Institute of Psychology, Chinese Academy of Sciences and the Nathan Kline Institute. | Zuo et al., 2014 |

| | | | | |
|---|---|---|---|---|
| Dallas Lifespan Brain Study | DLBS | https://fcon_1000.projects.nitrc.org/indi/retro/dlbs.html | We would like to thank the following individuals and research bodies for their continuing support of the study: Our scientific and support staff, who conduct the day-to-day operations and provide the long-term support necessary to keep the study running: Paula Abercrombie, Bela Bhatia, Gerard Bischof, Ph.D., Micaela Chan, Xi Chen, Arielle Click, Mark Diaz-Arrastia, Aaron Dostson, Linda Dubose, Patrick Evans, Victor Faner, Michelle Farrell, Blair Flicker, Jacqueline Gauer, Cassandra Hatt, Andy Hebrank, Marci Horn, Richard Innis, Caroline Janeway, Debby Kirchhevel, Mitchell Meltzer, April Norambuena, Heekyeong Park, Ph.D., Allison Parker, Jenny Rieck, Ph.D., Melissa Rundle, Ph.D., Prasanna Tamil, Nicole Tehrani, Erin Wooden. The Center for Vital Longevity, the University of Texas at Dallas, and the University of Texas Southwestern Medical Center, for sponsoring and providing the support and facilities needed to conduct the study. The National Institutes of Health and Aging, for their continuing financial and scientific support. AVID Radiopharmaceuticals, for providing the ligand used in the PET Imaging procedure. The Aging Mind Foundation and the Alzheimer's Association, for providing additional funding for this research. Finally, the Dallas Lifespan Brain Study could not have been accomplished without the help of our research participants. We appreciate their continued interest and participation in the study! | H. Lu et al., 2011 |
| 1000 Functional Connectomes | FCON1000 | https://fcon_1000.projects.nitrc.org/fcpClassic/FcpTable.html | Collected at 33 independent sites by J.J. Pekar, S.H. Mostofsky, S, Colcombe, Y.F. Zang, D. Marguiles, R.L. Buckner, M.J Low, B. Rypma, D.J. Madden, A.C. Evans, S.A.R.B. Rombouts, A. Villringer, S.J. Li, C.Sorg, V. Riedel, B. Biswal, M. Hampson, M.P. Milham, F.X. Castellanos, P. Williamson, M. Hoptman, V.J. Kiviniemi, J. Veijiola, S.M. Smith, C. Mackay, M. Greicius, G. Siegle, K. McMahon, B. Schlaggar, S. Petersen, C.P. Lin, H.S. Mayberg, C.S. Monk, R.D. Seidler, S.J. Peltier. | |
| Healthy Brain Network | HBN | https://data.healthybrainnetwork.org/ | We thank the Communications, Development, Finance, and Human Resource teams at the Child Mind Institute (past and present) for their endless support, as well as the CMI Executive Team and the Child Mind Institute Scientific Research Council for their guidance and critical feedback in the planning of the Healthy Brain Network; Judith Gardner and Bernard Karmel for assisting in the recruitment efforts; Tammy Vanderwaal and Uri Hasson for their | Alexander et al., 2017 |

| | | | |
|---|---|---|---|
| | | | consultation in the selection of movies for the natural viewing paradigms; Simon Kelly for his assistance with devising the EEG battery; Megan Horton for advising us to add the collection of baby teeth; Antonio Convit for information regarding assessments of body composition; Stan Colcombe for information regarding fitness assessments; and Michael Michaelides for advising us to add hair samples for metals. Additionally, we would like to thank Joan Kaufman and Ken Kobak for providing access to the newly developed computerized KSADS and Ted Satterthwaite for helpful comments on the manuscript during its preparation. We also acknowledge and thank Staten Island Borough President James Oddo, Staten Island Health and Wellness Director Dr. Ginny Mantello, and New York State Senator Andrew J. Lanza and his team (specifically William Matarazzo and Anthony Reinhart) for their guidance in developing strong partnerships throughout Staten Island, and their continued support of the project. We would also like to express our sincere gratitude to the mental health organizations, service providers, and clinicians across Staten Island, and NYC at large, who continue to work with our staff and refer participants to the project. Our sincere gratitude is extended to the participants and their families for their contributions to this project. The Healthy Brain Network and its collaborative initiatives are supported by philanthropic contributions from the following individuals, foundations and organizations: Margaret Bilotti; Brooklyn Nets; Agapi and Bruce Burkard; James Chang; Phyllis Green and Randolph Cowen; Grieve Family Fund; Susan Miller and Byron Grote; Sarah and Geoff Gund; George Hall; Jonathan M. Harris Family Foundation; Joseph P. Healey; The Hearst Foundations; Eve and Ross Jaffe; Howard & Irene Levine Family Foundation; Rachael and Marshall Levine; George and Nitzia Logothetis; Christine and Richard Mack; Julie Minskoff; Valerie Mnuchin; Morgan Stanley Foundation; Amy and John Phelan; Roberts Family Foundation; Jim and Linda Robinson Foundation, Inc.; Linda and Richard Schaps; Zibby Schwarzman; Abigail Pogrebin and David Shapiro; Stavros Niarchos Foundation; Preethi Krishna and Ram Sundaram; Amy and John Weinberg; Donors to the 2013 Child Advocacy Award Dinner Auction; Donors to the 2012 Brant Art Auction. |

| | | | | |
|---|---|---|---|---|
| Human Connectome Project | HCP | https://www.humanconnectome.org/ | Data were provided in part by the Human Connectome Project, MGH-USC Consortium (Principal Investigators: Bruce R. Rosen, Arthur W. Toga and Van Wedeen; U01MH093765) funded by the NIH Blueprint Initiative for Neuroscience Research grant; the National Institutes of Health grant P41EB015896; and the Instrumentation Grants S10RR023043, 1S10RR023401, 1S10RR019307. | Van Essen et al., 2013 |
| Amsterdam Open MRI Collection - ID1000 | ID1000 | https://openneuro.org/datasets/ds003097/versions/1.2.1 | We thank all research assistants and students who helped collecting the data of the three projects, Jasper Wijnen and Marco Teunisse for advice and guidance with respect to anonymization and GDPR-related concerns, and Jos Bloemers, Sennay Ghebeab, Adriaan Tuiten, Joram van Driel, Christian Olivers, Ilja Sligte, Sara Jahfari, Guido van Wingen, and Suzanne Oosterwijk for help with designing the paradigms, Marcus Spaan for technical support, and Franklin Feingold and Joe Wexler for help with uploading the datasets to Openneuro. | Snoek et al., 2021 |
| Information eXtraction from Images | IXI | https://brain-development.org/ixi-dataset/ | | |
| Minimal Interval Resonance Imaging in Alzheimer's Disease | MIRIAD | https://www.ucl.ac.uk/drc/research-clinical-trials/minimal-interval-resonance-imaging-alzheimers-disease-miriad | Data used in the preparation of this article were obtained from the MIRIAD database. The MIRIAD investigators did not participate in analysis or writing of this report. The MIRIAD dataset is made available through the support of the UK Alzheimer's Society (Grant RF116). The original data collection was funded through an unrestricted educational grant from GlaxoSmithKline (Grant 6GKC). | Malone et al., 2013 |
| Max Planck Institut Leipzig Mind-Brain-Body Dataset | MPI-LEMON | https://fcon_1000.projects.nitrc.org/indi/retro/MPI_LEMON.html | We thank all participants who volunteered to participate in our study. Moreover, we thank Elizabeth Kelly for proofreading the manuscript and Heike Schmidt-Duderstedt for editing tables and figures. | Babayan et al., 2019; Mendes et al., 2019 |
| Parkinson's Disease Datasets (NEUROCON) | NEUROCON | https://fcon_1000.projects.nitrc.org/indi/retro/parkinsons.html | This work was partially supported by the NEUROCON project (84/2012), financed by UEFISCDI. | Badea et al., 2017 |
| National Institute of Mental Health - Healthy Research Volunteer dataset | NIMH | https://openneuro.org/datasets/ds005752/versions/2.1.0 | We thank the NIMH Office of the Clinical Director, the outpatient behavioral health clinic and NMR center for providing support for the data collection. This work utilized the computational resources of the NIH HPC Biowulf cluster http://hpc.nih.gov. We thank Sil van der Woerd for graciously allowing us to use his film as a behavioral task. In addition, we thank the subjects who generously contributed their data to this project. | Nugent et al., 2022 |
| Enhanced Nathan Kline Institute - Rockland Sample | NKI-RS | https://fcon_1000.projects.nitrc.org/indi/enhanced/ | We would like to thank Lawrence Maayan for his key role in the design and execution of the pilot NKI-RS | Nooner et al., 2012; Tobe et al., 2022 |

| | | | | |
|---|---|---|---|---|
| Open Access Series of Imaging Studies 3 | OASIS3 | https://sites.wustl.edu/oasisbrains/home/oasis-3/ | Data were provided in part by OASIS-3: Longitudinal Multimodal Neuroimaging: Principal Investigators: T. Benzinger, D. Marcus, J. Morris; NIH P30 AG066444, P50 AG00561, P30 NS09857781, P01 AG026276, P01 AG003991, R01 AG043434, UL1 TR000448, R01 EB009352. AV-45 doses were provided by Avid Radiopharmaceuticals, a wholly owned subsidiary of Eli Lilly. | LaMontagne et al., 2019 |
| Pediatric Imaging, Neurocognition, and Genetics | PING | https://chd.ucsd.edu/research/ping-study.html | Data used in the preparation of this article were obtained from the Pediatric Imaging, Neurocognition and Genetics (PING) Study database (www.chd.ucsd.edu/research/ping-study.html, now shared through the NIMH Data Archive (NDA)). PING was a multisite, cross-sectional study that recruited more than 1,700 participants aged 3 to 20 years. The study was supported by award number RC2DA029475 from the National Institute on Drug Abuse with additional support for data sharing provided by the Eunice Kennedy Shriver National Institute of Child Health & Human Development under award number R01HD061414. A list of participating sites and study investigators can be found at https://ping-dataportal.ucsd.edu/sharing/Authors10222012.pdf. PING investigators designed and implemented the study and/or provided data but did not necessarily participate in analysis or writing of this report. This publication is solely the responsibility of the authors and does not necessarily represent the views of the National Institutes of Health or PING investigators. | Jernigan et al., 2016 |
| Amsterdam Open MRI Collection - PIOP1 | PIOP1 | https://openneuro.org/datasets/ds002785/versions/2.0.0 | We thank all research assistants and students who helped collecting the data of the three projects, Jasper Wijnen and Marco Teunisse for advice and guidance with respect to anonymization and GDPR-related concerns, and Jos Bloemers, Sennay Ghebeab, Adriaan Tuiten, Joram van Driel, Christian Olivers, Ilja Sligte, Sara Jahfari, Guido van Wingen, and Suzanne Oosterwijk for help with designing the paradigms, Marcus Spaan for technical support, and Franklin Feingold and Joe Wexler for help with uploading the datasets to Openneuro. | Snoek et al., 2021 |
| Amsterdam Open MRI Collection - PIOP2 | PIOP2 | https://openneuro.org/datasets/ds002790/versions/2.0.0 | We thank all research assistants and students who helped collecting the data of the three projects, Jasper Wijnen and Marco Teunisse for advice and guidance with respect to anonymization and GDPR-related concerns, and Jos Bloemers, Sennay Ghebeab, Adriaan Tuiten, Joram van Driel, Christian Olivers, Ilja Sligte, Sara Jahfari, Guido van Wingen, and | Snoek et al., 2021 |

| | | | | |
|---|---|---|---|---|
| | | | Suzanne Oosterwijk for help with designing the paradigms, Marcus Spaan for technical support, and Franklin Feingold and Joe Wexler for help with uploading the datasets to Openneuro. | |
| Philadelphia Neurodevelopmental Cohort | PNC | https://www.med.upenn.edu/bbl/philadelphianeurodevelopmentalcohort.html | Support for the collection of the data sets was provided by grant RC2MH089983 awarded to R. Gur and RC2MH089924 awarded to H. Hakonarson. | Satterthwaite et al., 2016; Satterthwaite et al., 2014 |
| Parkinson's Progression Markers Initiative | PPMI | https://www.ppmi-info.org/ | This analysis used data openly available from PPMI. PPMI – a public-private partnership – is funded by the Michael J. Fox Foundation for Parkinson's Research and funding partners, including 4D Pharma, Abbvie, AcureX, Allergan, Amathus Therapeutics, Aligning Science Across Parkinson's, AskBio, Avid Radiopharmaceuticals, BIAL, BioArctic, Biogen, Biohaven, BioLegend, BlueRock Therapeutics, Bristol-Myers Squibb, Calico Labs, Capsida Biotherapeutics, Celgene, Cerevel Therapeutics, Coave Therapeutics, DaCapo Brainscience, Denali, Edmond J. Safra Foundation, Eli Lilly, Gain Therapeutics, GE HealthCare, Genentech, GSK, Golub Capital, Handl Therapeutics, Insitro, Jazz Pharmaceuticals, Johnson & Johnson Innovative Medicine, Lundbeck, Merck, Meso Scale Discovery, Mission Therapeutics, Neurocrine Biosciences, Neuron23, Neuropore, Pfizer, Piramal, Prevail Therapeutics, Roche, Sanofi, Servier, Sun Pharma Advanced Research Company, Takeda, Teva, UCB, Vanqua Bio, Verily, Voyager. | Marek et al., 2018 |
| Queensland Twin Adolescent Brain | QTAB | https://openneuro.org/datasets/ds004146/versions/1.0.4 | We are forever grateful to the twins and their families for their willingness to participate in our studies. We thank Liza van Eijk, Victoria O'Callaghan, Islay Davies, Ethan Campi, Kimberley Huang, Eleanor Roga, Michael Day, Aiman Al-Najjar, Zoie Nott, Tom Shaw, Nicole Atcheson, and Sarah Daniel for data acquisition. We thank Naomi Wray for funding the collection of metabolic samples, including detailed dietary data, Ian Hickie and Kathleen Merikangas for funding and support of actigraphy data, and Sarah Medland and ENIGMA GWAS for funding genotyping. Special thanks to Anjali Henders, Leanne Wallace, Lorelle Nunn and the many laboratory assistants at the Human Studies Unit (part of the Program in Complex Trait Genomics based at the Institute of Molecular Bioscience, University of Queensland) for the processing and storage of biological samples. Thanks also to Julie Henry for helpful discussion of social cognition measures. The QTAB project was funded by the National Health and Medical Research Council (NHMRC), Australia | Strike et al., 2023 |

| | | | | |
|---|---|---|---|---|
| | | | (Project Grant ID: 1078756 to MJW), the Queensland Brain Institute, University of Queensland, and with the assistance of resources from the Centre for Advanced Imaging and the Queensland Cyber Infrastructure Foundation, University of Queensland. We acknowledge the Queensland Twin Registry (QTwin) (https://www.qimrberghofer.edu.au/study/queensland-twin-registry-study) for generously sharing database information for recruitment. Recruitment was further facilitated through access to Twins Research Australia, a national resource supported by a Centre of Research Excellence Grant (ID: 1079102) from the NHMRC. Lastly, we thank the many researchers worldwide for providing access to their assessments. | |
| Queensland Twin IMaging | QTIM | https://openneuro.org/datasets/ds004169/versions/1.0.6 | We are forever grateful to the twins and siblings for their willingness to participate in our studies. We thank Marlene Grace and Ann Eldridge for participant recruitment; Kerrie McAloney for study co-ordination; Kori Johnson, Aaron Quiggle, Natalie Garden, Matthew Meredith, Peter Hobden, Kate Borg, Aiman Al Najjar and Anita Burns for data acquisition; David Butler and Daniel Park for IT-support. | Strike et al., 2019 |
| Southwest University Adult Lifespan Dataset | SALD | https://fcon_1000.projects.nitrc.org/indi/retro/sald.html | This data repository was supported by the National Natural Science Foundation of China (31470981; 31571137; 31500885), National Outstanding young people plan, the Program for the Top Young Talents by Chongqing, the Fundamental Research Funds for the Central Universities (SWU1509383, SWU1509451, SWU1609177), Natural Science Foundation of Chongqing (cstc2015jcyjA10106), Fok Ying Tung Education Foundation (151023) , General Financial Grant from the China Postdoctoral Science Foundation (2015M572423, 2015M580767), Special Funds from the Chongqing Postdoctoral Science Foundation (Xm2015037, Xm2016044), Key research for Humanities and social sciences of Ministry of Education (14JJD880009). | Wei et al., 2018 |
| SchizConnect | SCZ-C | https://schizconnect.org/ | Data used in preparation of this article were obtained from the SchizConnect (http://schizconnect.org) database. As such, the investigators within SchizConnect contributed to the design and implementation of SchizConnect and/or provided data but did not participate in analysis or writing of this report. Data collection and sharing for this project was funded by NIMH cooperative agreement 1U01 MH097435 SCHIZCONNECT1 comprised BrainGluSchi, COBRE and MCIC samples (COINS). SCHIZCONNECT2 comprised | Ambite et al., 2015; Bustillo et al., 2017; Çetin et al., 2014; Gollub et al., 2013; Kogan et al., 2016; L. Wang et al., 2016 |

| | | | | |
|---|---|---|---|---|
| | | | NUSDAST and NUNDA samples. Duplicate subjects in different sources were excluded. The respective samples were supported by the following grants: BrainGluSchi: NIMH R01MH084898-01A1. COBRE: 5P20RR021938 /P20GM103472 from the NIH to Dr. Vince Calhoun. MCIC: Department of Energy under Award Number DEFG02-08ER64581. NUSDAST: NIMH Grant 1R01 MH084803. NUNDA: MH056584. | |
| Southwest University Longitudinal Imaging Multimodal dataset | SLIM | https://fcon_1000.projects.nitrc.org/indi/retro/southwestuni_qiu_index.html | We are grateful to all of graduate students who contributed their time and wisdom to this data repository, including but not limited to Xue Du, Kang cheng Wang, Jiangzhou Sun, Qunling Chen and Wei Liu. We are also thankful for assistance from Michael Milham and David O'Connor (Child Mind Institute) in constructing the webpage. This data repository was supported by: The National Natural Science Foundation of China (31271087; 31470981; 31571137; 31500885), National Outstanding young people planthe Program for the Top Young Talents by Chongqing, the Fundamental Research Funds for the Central Universities (SWU1509383,SWU1509451), Natural Science Foundation of Chongqing (cstc2015jcyjA10106), Fok Ying Tung Education Foundation (151023), General Financial Grant from the China Postdoctoral Science Foundation (2015M572423, 2015M580767), Special Funds from the Chongqing Postdoctoral Science Foundation (Xm2015037), Key research for Humanities and social sciences of Ministry of Education(14JJD880009). | Y. Wang et al., 2014 |
| StrokeMRI | StrokeMRI | Authors | Supported by the Research Council of Norway (249795, 248238), the South-Eastern Norway Regional Health Authority (2014097, 2015044, 2015073, 2016083), and the Norwegian ExtraFoundation for Health and Rehabilitation (2015/FO5146). | Dørum et al., 2016 |
| Thematically Organized Psychosis | TOP | Authors | The work was funded by the Research Council of Norway (213837, 223273, 204966/F20, 213694, 229129, 249795/F20, 248778), the South-Eastern Norway Regional Health Authority (2013-123, 2014-097, 2015-073, 2017-112) and Stiftelsen Kristian Gerhard Jebsen. | Brandt et al., 2015; Kaufmann et al., 2015; Skåtun et al., 2016 |
| Parkinson's Disease Datasets (Tao-Wu) | Tao-Wu | https://fcon_1000.projects.nitrc.org/indi/retro/parkinsons.html | This work was partially supported by the NEUROCON project (84/2012), financed by UEFISCDI. | Badea et al., 2017 |
| UK Biobank | UKBB | https://www.ukbiobank.ac.uk/ | This research has been conducted using the UK Biobank Resource (access code 27412). | Sudlow et al., 2015 |
| | ds000119 | https://openneuro.org/datasets/ds000119/versions/00001 | We thank Mark McAvoy and Abraham Snyder for support and development of functional data analysis procedures. Enami Yasui provided | Velanova et al., 2008 |

| | | | | |
|---|---|---|---|---|
| | | | assistance with data collection. National Institutes of Mental Health (NIMH RO1 MH067924). | |
| | ds000171 | https://openneuro.org/datasets/ds000171/versions/00001 | The authors wish to acknowledge Trisha Patrician and Natalie Stroupe for their assistance with screening of participants, and Allan Schmitt and Franklin Hunsinger for their role in collecting the MR data. | Lepping et al., 2016 |
| | ds000202 | https://openneuro.org/datasets/ds000202/versions/00001 | | Van Schuerbeek et al., 2016 |
| | ds000222 | https://openneuro.org/datasets/ds000222/versions/1.0.1 | Citation of FitzGerald et al. Sequential inference as a mode of cognition and its correlates in fronto-parietal and hippocampal brain regions. PLoS Computational Biology (2017) This data was obtained from the OpenfMRI database. Its accession number is ds000222. | FitzGerald et al., 2017 |
| | ds000245 | https://openneuro.org/datasets/ds000245/versions/00001 | This data was obtained from the OpenfMRI database. Its accession number is ds000245. | Yoneyama et al., 2018 |
| | ds002424 | https://openneuro.org/datasets/ds002424/versions/1.2.0 | This research was supported by NIH grant R21-MH080820 to J. R. Booth, and the Northwestern University Human Cognition T32-NS047987 NIH training grant to R. Hammer. | Lytle et al., 2020 |
| | ds003592 | https://openneuro.org/datasets/ds003592/versions/1.0.9 | | Spreng et al., 2022 |
| | ds004302 | https://openneuro.org/datasets/ds004302/versions/1.0.1 | The authors wish to thank Dr Aniol Santo-Angles for his work and support in data acquisition and analysis of this project. This work was supported by the CIBERSAM and the Catalonian Government (2017SGR01271 to EP-C and 2017SGR1265 to WH). Also by a grant from the Plan Nacional de I+D+i 2013–2016: Juan de la Cierva-formación contract (FJCI-2015-25278 to PF-C) and two projects from the Ministerio de Ciencia, Innovación y Universidades (MCIU) y la Agencia Estatal de Investigación (AEI), FFI2016-77647-C2-2-P to WH and PS-P and PID2019-110120RBI00/AEI/10.13039/501100011033 to WH; and by the Instituto de Salud Carlos III, co-funded by European Union (ERDF/ESF, "Investing in your future"): Miguel Servet Research contracts (CPII13/00018 to RS and MS10/00596 to EP-C), Sara Borrell contract (CD19/00149 to PF-C) and Research Project Grants (PI18/00880 to PM). The funders had no role in study design; in the collection, analysis and interpretation of data; in the writing of the report; or in the decision to submit the article for publication. | Soler-Vidal et al., 2022 |
| | ds004392 | https://openneuro.org/datasets/ds004392/versions/1.0.0 | The authors wish to thank Christine Martin, Sarah Rogers, and Abigail Simpson for their help with participant recruitment and | Wylie et al., 2023 |

| | | | |
|---|---|---|---|
| | | | neurophysiological testing administration. Funding: NIH Grant 1K02NS080885-01A1, NIH Grant 1R21NS093266-01A1, NIH Grant K01 AT009894-01, Michael J. Fox Foundation for Parkinson's Research Grant Number 10879. |

Supplementary Table 1: An overview of the origins and acknowledgements for the data sources used in the study

| Dataset characteristics | | | | | Usage in the study | |
|---|---|---|---|---|---|---|
| **Name** | **Images** | **Participants** | **Ages** | **Females** | **Pretraining** | **Downstream** |
| ABCD | 17,166 | 11,660 | 10 ± 1 | 47% | ✓ | |
| ABIDE-I | 973 | 973 | 17 ± 8 | 16% | | ✓ |
| ABIDE-II | 876 | 876 | 15 ± 10 | 22% | | ✓ |
| ADHD200 | 804 | 804 | 12 ± 3 | 38% | | ✓ |
| ADNI | 20,670 | 2729 | 75 ± 7 | 45% | ✓ | ✓ |
| AIBL | 906 | 590 | 74 ± 7 | 54% | | ✓ |
| Beijing | 180 | 180 | 21 ± 2 | 59% | ✓ | |
| BRAINMINT | 681 | 674 | 18 ± 3 | 72% | | ✓ |
| Cam-CAN | 653 | 653 | 55 ± 19 | 51% | ✓ | |
| CoRR | 2,275 | 1,156 | 26 ± 16 | 51% | ✓ | |
| DLBS | 315 | 315 | 55 ± 20 | 63% | ✓ | |
| FCON1000 | 853 | 836 | 29 ± 15 | 54% | ✓ | |
| HBN | 1,369 | 1,365 | 10 ± 4 | 35% | | ✓ |
| HCP | 1,113 | 1,113 | 29 ± 4 | 54% | ✓ | |
| ID1000 | 2,441 | 819 | 23 ± 2 | 54% | ✓ | |
| IXI | 534 | 534 | 48 ± 16 | 55% | ✓ | |
| MIRIAD | 324 | 69 | 69 ± 7 | 58% | | ✓ |
| MPI-LEMON | 227 | 227 | 39 ± 20 | 36% | ✓ | |
| NEUROCON | 43 | 43 | 68 ± 11 | 51% | | ✓ |
| NIMH | 249 | 155 | 34 ± 13 | 67% | ✓ | |
| NKI-RS | 867 | 664 | 35 ± 21 | 57% | ✓ | |
| OASIS3 | 2,773 | 1,062 | 67 ± 9 | 57% | | ✓ |
| PING | 1,177 | 1,176 | 12 ± 5 | 48% | ✓ | |
| PIOP1 | 209 | 209 | 22 ± 2 | 57% | ✓ | |
| PIOP2 | 224 | 224 | 22 ± 2 | 57% | ✓ | |
| PNC | 835 | 835 | 15 ± 3 | 54% | ✓ | |
| PPMI | 1,176 | 784 | 64 ± 9 | 41% | | ✓ |
| QTAB | 718 | 414 | 12 ± 2 | 50% | ✓ | |
| QTIM | 1,341 | 1,201 | 21 ± 4 | 61% | ✓ | |
| SALD | 494 | 494 | 45 ± 17 | 62% | ✓ | |
| SCZ-C | 710 | 710 | 34 ± 12 | 33% | | ✓ |
| SLIM | 1,023 | 575 | 20 ± 1 | 55% | ✓ | |
| StrokeMRI | 588 | 341 | 61 ± 14 | 62% | | ✓ |
| TOP | 2,702 | 2,222 | 32 ± 10 | 45% | | ✓ |
| Tao-Wu | 40 | 40 | 65 ± 5 | 42% | | ✓ |
| UKBB | 45,891 | 45,891 | 65 ± 8 | 52% | ✓ | |
| ds000119 | 73 | 73 | 16 ± 5 | 59% | ✓ | |
| ds000171 | 39 | 39 | 31 ± 12 | 56% | | ✓ |
| ds000202 | 95 | 95 | 22 ± 3 | 100% | ✓ | |
| ds000222 | 79 | 79 | 44 ± 20 | 52% | ✓ | |
| ds000245 | 45 | 45 | 66 ± 7 | 56% | | ✓ |
| ds002424 | 77 | 77 | 10 ± 1 | 18% | | ✓ |
| ds003592 | 301 | 301 | 41 ± 23 | 56% | ✓ | |
| ds004302 | 71 | 71 | 42 ± 12 | 24% | | ✓ |
| ds004392 | 57 | 57 | 70 ± 7 | 33% | | ✓ |

Supplementary Table 2: An overview of the high-level characteristics of the datasets used in the study, and what part of the process they were utilised for.

| Cohort | Field identifier | Description |
|---|---|---|
| ABCD | nihtbx_fluidcomp_uncorrected | |
| ID1000 | IST_fluid | IST fluid intelligence subscale |
| UKBB | 20016 | This is a simple unweighted sum of the number of correct answers given to the 13 fluid intelligence questions. Participants who did not answer all of the questions within the allotted 2 minute limit are scored as zero for each of the unattempted questions. |

Supplementary Table 3: The columns from the various cohorts that were used to derive the unified fluid intelligence score

| Cohort | Field identifier | Description |
|---|---|---|
| ID1000 | NEO_N | Neuroticism scale (sum score) |
| PIOP1 | NEO_N | Neuroticism scale (sum score) |
| PIOP2 | NEO_N | Neuroticism scale (sum score) |
| UKBB | 20127 | This is an externally derived summary score of neuroticism, based on 12 neurotic behaviour domains as reported from fields 1920, 1930, 1940, 1950, 1960, 1970, 1980, 1990, 2000, 2010, 2020 and 2030 from the touchscreen questionnaire at baseline. Participants were assessed for twelve domains of neurotic behaviours via the touchscreen questionnaire. Questions included:. This field summarises the number of Yes answers across these twelve questions into a single integer score for each participant. This derived data field has come from Professor Jill Pell from the Institute of Health & Wellbeing, University of Glasgow.<br>• Does your mood often go up and down?<br>• Do you ever feel 'just miserable' for no reason?<br>• Are you an irritable person?<br>• Are your feelings easily hurt?<br>• Do you often feel 'fed-up'?<br>• Would you call yourself a nervous person?<br>• Are you a worrier?<br>• Would you call yourself tense or 'highly strung'?<br>• Do you worry too long after an embarrassing experience?<br>• Do you suffer from 'nerves'?<br>• Do you often feel lonely?<br>• Are you often troubled by feelings of guilt?<br>Participants could answer Yes, No, Do not know or Prefer not to answer. This field summarises the number of Yes answers across these twelve questions into a single integer score for each participant. This derived data field has come from Professor Jill Pell from the Institute of Health & Wellbeing, University of Glasgow. |

Supplementary Table 4: The columns from the various cohorts that were used to derive the unified neuroticism score

| | Age | Sex | Handedness | Body mass index | Neuroticism | Fluid intelligence |
|---|---|---|---|---|---|---|
| Age | 1 | 0 | 0.02 | 0.52 | -0.1 | -0.04 |
| Sex | 0 | 1 | 0.03 | 0.04 | -0.15 | 0.04 |
| Handedness | -0.02 | 0.03 | 1 | 0 | 0.03 | -0.03 |
| Body mass index | 0.52 | 0.04 | 0 | 1 | -0.03 | -0.05 |
| Neuroticism | -0.1 | -0.15 | 0.03 | -0.03 | 1 | -0.04 |
| Fluid intelligence | -0.04 | 0.04 | -0.03 | -0.05 | -0.04 | 1 |

Supplementary Table 5: Correlation between the targets used for multi-task pretraining. Correlation between continuous variables were measured using the Spearman rank coefficient, between continuous and binary variables using the point biserial coefficient, and between binary variables using the Matthews coefficient.
